# Interdental and subgingival microbiota may affect the tongue microbial ecology and oral malodour in health, gingivitis and periodontitis

**DOI:** 10.1101/2021.04.29.21256204

**Authors:** Abish S. Stephen, Narinder Dhadwal, Vamshidhar Nagala, Cecilia Gonzales-Marin, David G. Gillam, David J. Bradshaw, Gary R. Burnett, Robert P. Allaker

## Abstract

**Background and Objective:** Oral malodour is often observed in gingivitis and chronic periodontitis patients, and the tongue microbiota is thought to play a major role in malodorous gas production, including Volatile Sulfur Compounds (VSCs) such as hydrogen sulfide (H_2_S) and methanethiol (CH_3_SH). This study aimed to examine the link between the presence of VSCs in mouth air (as a marker of oral malodour) and the oral bacterial ecology in the tongue and periodontal niches of healthy, gingivitis and periodontitis patients.

**Methods:** Participants were clinically assessed using plaque index, bleeding on probing (BOP) and periodontal probing depths, and VSC concentrations in their oral cavity measured using a portable gas chromatograph. Tongue scrapings, subgingival and interdental plaque were collected from healthy individuals (n=22), and those with gingivitis (n=14) or chronic periodontitis (n=15). The bacterial 16S rRNA gene region V3-V4 in these samples was sequenced and the sequences analysed using the Minimum Entropy Decomposition pipeline.

**Results:** Elevated VSC concentrations and CH_3_SH:H_2_S were observed in periodontitis compared to health. Significant ecological differences were observed in the tongue microbiota of healthy subjects with high plaque scores compared to low plaque scores, suggesting a possible connection between the microbiota of the tongue and the periodontium and that key dysbiotic changes may be initiated in the clinically healthy who have higher dental plaque accumulation. Greater subgingival bacterial diversity was positively associated with H_2_S in mouth air. Periodontopathic bacteria known to be prolific VSC producers increased in abundance on the tongue associated with increased bleeding on probing (BOP) and total percentage of periodontal pockets >6mm, supporting the suggestion that the tongue may become a reservoir for periodontopathogens.

**Conclusion:** This study highlights the importance of the periodontal microbiota in malodour and has detected dysbiotic changes in the tongue microbiota in periodontitis.

## INTRODUCTION

Intra-oral halitosis is often observed in patients with periodontitis, and, of the volatile sulfur compounds detected in the breath in association with this condition, an elevated methanethiol to hydrogen sulfide ratio has been observed in patients compared to healthy controls [1,2]. The severity of oral malodour and methanethiol to hydrogen sulfide ratio (CH_3_SH: H_2_S) is also positively correlated with the clinical parameters of periodontitis in individuals complaining of oral malodour [3,4]. Ecological population dynamics involving different bacterial taxa have been known to play a role in the aetiopathogenesis of both gingivitis and chronic periodontitis, with studies utilizing sequencing methodologies identifying a number of bacterial taxa responsible for altering the overall ecology of the oral soft and hard tissue niches in the different disease states [5-8]. Of these niches, the tongue biofilm is considered the primary source of oral malodour in healthy individuals, with some clinical studies suggesting that plaque accumulation in the periodontal niches such as the subgingival and supragingival aspects of teeth may play a role in periodontitis-associated oral malodour [9-11]. In addition, an interventional study has reported significant reductions in H_2_S and CH_3_SH, in the breath of patients undergoing periodontal treatment as opposed to just tongue cleaning [11,12].

Current surveys employing next generation bacterial rRNA gene sequencing to study oral malodour associated microbiota, have only investigated the tongue and saliva, identifying some important bacterial taxa that may be associated with oral malodour in healthy individuals [13-15]. An early study that used pyrosequencing of tongue biofilm samples of healthy individuals reported interpersonal variation in community structure to be smaller than when associated with VSC concentrations, with the genera *Prevotella* and *Leptotrichia* to be positively associated with higher H_2_S concentrations in the breath [13]. Another study that explored the salivary microbiome in individuals grouped as high H_2_S or CH_3_SH found genera such as *Neisseria, Fusobacterium, Porphyromonas* and *Absconditabacteria* (SR1) in higher proportions in the individuals with high concentrations of H_2_S in mouth air [15]. A recent study of the tongue microbiome of intra-oral halitosis also suggested no significant difference in overall community structure but some differences in several OTU abundance [14].

Studies exploring the microbiota involved in periodontitis typically do not include sampling the tongue, but separate studies have identified taxa that may be involved in disease progression, associated with a particular stage of periodontitis in the periodontal niches or tongue [16,17]. However, there appear to be no available studies using sequencing approaches to investigate the tongue and periodontal microbiota within the same individuals in the context of disease and oral malodour status. This study, therefore, aimed to explore how tongue and periodontal microbiota vary in individuals with gingivitis, periodontitis, and periodontal health, with respect to intra-oral malodour and clinical indices of periodontitis. In this paper, we first explore if alpha diversities of the tongue and periodontal microbiota are associated with breath VSCs and clinical parameters such as probing depth, bleeding on probing and plaque index. We then analyse if either oral malodour or clinical diagnosis (health, gingivitis, periodontitis) can better explain the observed variation in the beta diversity of tongue, subgingival and interdental microbiota. Finally, we identify the microbial taxa responsible for the ecological shifts observed on the tongue and periodontal niches in association with oral malodour and periodontitis. It was hypothesised in accordance with the literature that there may be no overall difference in the tongue microbiota of individuals with or without oral malodour, but there may be differences at the species level, particularly in relation to periodontitis.

## MATERIALS AND METHODS

### Study design, clinical assessments and participants

This was a case control study consisting of three groups of participants (i) health (ii) gingivitis (iii) chronic periodontitis, recruited between February 2013 and May 2015 from Queen Mary University of London and Royal London Dental Hospital (Barts Health NHS Trust) based on clinically determined oral health parameters. Ethical approval for the study protocol was obtained from the London Central Research Ethics Committee (12/LO/1611) and conformed to STROBE guidelines for human case control studies. All study procedures were in accordance with the Declaration of Helsinki. Written informed consent was obtained from eligible participants aged 18 to 70 years, with at least 20 teeth, provided the following exclusion criteria were not met: presence of systemic diseases and/or respiratory infections, women intending to be or are pregnant, smokers, use of antibiotics and/or anti-inflammatory medication 4 weeks prior to visit, use of medications resulting in xerostomia, periodontal treatment in the previous 3 months, denture wearers.

Participants were screened and recruited into the different groups based on the following clinical oral parameters: (i) Probing depth (PD - full mouth manual probing of six sites around all teeth present. (ii) Percentage of sites bleeding on probing (BOP) - bleeding observed in sites up to 20s after probing. The healthy group consisted of individuals with no observable carious lesions or periodontitis with no more than four periodontal pockets of a maximum depth of 4mm excluding third molars and ≤20% of sites BOP. This group was further stratified based on the observed plaque coverage [29]: ≤30% PI (low plaque) and >30% PI (high plaque). Individuals with generalised gingival inflammation, >20% of sites BOP, >30% PI and no obvious signs of periodontitis such as generalised periodontal pocketing were recruited to the gingivitis group. The chronic periodontitis group consisted of patients with radiographic evidence of bone loss, at least two PDs ≥6mm excluding third molars and >2mm Clinical Attachment Loss (CAL). The measured clinical parameters for the different cohorts are listed in Table 1.

**Table 1.** **Table listing demographic and breath data in the study population. Group comparisons that yielded statistical significance (p<0.05) are indicated with numbers in the last column (1=low plaque health vs high plaque health, 2=low plaque health vs gingivitis, 3=low plaque health vs periodontitis, 4=high plaque health vs gingivitis, 5=high plaque health vs periodontitis, 6=gingivitis vs periodontitis). †Fisher’s exact test.**

| Group (n) | Low plaque health (11) | High plaque health (11) | Gingivitis (15) | Periodontitis (14) | Statistically significant? |
| --- | --- | --- | --- | --- | --- |
| Mean age in years (SD) | 32.1 ± 6.8 | 32 ± 6.8 | 32.1 ± 6.7 | 41.4 ± 6.2 | 5, 6 |
| Male: Female | 3:8 | 5:6 | 5:8 | 5:9 | ns† |
| Mean plaque index (%) | 19.1 ± 6.8 | 47.9 ± 14.9 | 42.6 ± 22.1 | 49.0 ± 22.6 | 1,2,3 |
| Mean bleeding on probing (%) | 8.4 ± 6.7 | 12.5 ± 4.2 | 26.1 ± 13.7 | 28.0 ± 21.9 | 2,3 |
| Mean probing depths ≥4mm (%) | 1.3 ± 1.1 | 0.6 ± 0.5 | 7.1 ± 4.4 | 30.3 ± 14.3 | 3,5,6 |
| Mean probing depths ≥5mm (%) | 0 | 0 | 2.7 ± 2.2 | 23.7 ± 12.5 | 3,5,6 |
| Mean probing depths ≥6mm (%) | 0 | 0 | 0.3 ± 0.5 | 10.0 ± 6.0 | 3,5,6 |
| Mean [H<sub>2</sub>S] ppb (SD) | 132 (164) | 90 (101) | 165 (259) | 323 (582) | ns |
| Mean [CH<sub>3</sub>SH] ppb (SD) | 7 (16) | 22 (36) | 23 (53) | 63 (139) | ns |
| Mean CH<sub>3</sub>SH: H<sub>2</sub>S (SD) | 0.025 (0.047) | 0.31 (0.52) | 0.17 (0.27) | 0.12 (0.16) | 3 |
| Malodour prevalence (%) | 27.3% | 27.3% | 33.3% | 35.7% | ns† |

### Clinical protocol and sample collection

Participants were instructed not to eat or to perform oral hygiene for at least 2 hours before the visit. Participant restrictions included avoiding malodorous foods such as garlic, onion or coffee at least 12 hours prior to visit. The clinical protocol and sample collection sequence was carried out as shown in the Supplement.

#### Breath Sampling and Analysis

A previously detailed method [30] was used to collect breath samples. Briefly, participants were seated upright and breathed through their nose (mouth closed) for approximately one minute. A sterile mouthpiece (Fluorinated Ethylene Propylene tubing, Cole-Parmer, UK) with an attached 20 ml syringe (PolyTetraFluoroEthylene NORM-JECT, Henke-Sass Wolf, Germany) was then inserted into the mouth, ensuring that the mouthpiece reached the posterior half of the tongue. The participant held the mouthpiece with the lips closed (but not sealed). Approximately 15 ml of mouth air was withdrawn, as the participant took a deep inhalation through the nose and held for 5 seconds. A 1 ml aliquot of the breath sample was withdrawn from the 20 ml syringe and immediately injected into a portable gas chromatograph (Oral Chroma CHM-1, Abimedical Corp, Japan), to measure the concentration of hydrogen sulfide and methanethiol. The data output of the Oral Chroma in parts per billion (ppb) was recorded in the included data manager software (Abimedical Corp) and used for further data analysis. Samples that contained H_2_S >100ppb and/or CH_3_SH >50ppb were considered to be malodorous and used to calculate the malodour prevalence rates in each cohort [31].

#### Tongue biofilm, Subgingival and Interdental plaque sampling

The tongue biofilm was sampled by scraping a Whatman Omniswab (General Electric Healthcare, UK) three times along the tongue dorsum, from as close to the circumvallate papillae as possible towards the tip, with the swab held perpendicular to the lingual plane [32]. The Omniswab pad was then ejected into Tris-EDTA (TE) buffer (pH 7.2; Sigma-Aldrich, UK).

The tooth in each quadrant with the deepest pocket (excluding third molars), was sampled for subgingival plaque, after removal of supragingival plaque using Microbrush (Patterson Dental Supply Inc, USA). In case of quadrants with probing depths ≤3mm, a tooth was picked at random. Subgingival plaque samples were collected by inserting sterile absorbent paper points (Sybron Dental Specialities) in the deepest pockets in the buccal and lingual sites in each tooth for approximately 10 seconds [33]. Samples from all four teeth were pooled into TE buffer (Sigma).

Interdental plaque was collected from the mesial and distal aspects of all four teeth supragingivally, using an interdental brush [34] (Pink Tepe brush; Tepe Oral Hygiene Products, Sweden) and the brush stored immediately in TE buffer (Sigma).

### DNA extraction and sequencing

The plaque and tongue samples were disrupted from their respective sampling devices by transferring 3 to 5 sterile glass beads into the sample containing tubes and vortexing vigorously for 20s. DNA extraction was carried out on the plaque samples following the protocol recommended by Human Oral Microbiome Identification by Next Generation Sequencing (HOMINGS) using the MasterPure Gram-positive DNA purification kit (Epicentre, Madison, USA). After screening for quality, the samples were sequenced at the Forsyth Institute (USA) using the method described elsewhere [35]. Briefly, custom 341F-806R primers were used to amplify the bacterial 16S rRNA gene (V3-V4 region) in the samples and the libraries sequenced using the multiplexed, barcoded 250bp paired-end read methodology in the Illumina Miseq platform (Illumina, USA).

#### Sequence analysis

The raw Illumina reads were quality filtered, trimmed and merged with minimum overlap of 70 bases, Q30 check, a Q-score >20 for ambiguous bases recovered in the overlapping region and up to 2 ambiguous bases allowed in the overlap [36, 37]. Minimum Entropy Decomposition (MED) was used to partition these sequences into nodes at 1-nt resolution with the minimum substantive abundance parameter at 240 [38]. A BLAST search of the HOMD (v14.5) [39] and NCBI bacterial 16S databases classified representative sequences of all MED nodes, which was used to map the nodes to the Human Microbial Taxa (HMT) by percent identities. Alpha Diversity was calculated using the PAST software package (v3.22) [40] on the sequence abundance data of the nodes that collapsed to HMT. The raw sequence data are deposited in the NCBI Sequence Read Archive (BioProject ID: PRJNA649803).

### Multivariate analysis and statistical methods

Multivariate ordination and statistical analysis of the HOMINGS and MED sequence abundance data were performed using the PAST software package (v3.22). Differences in composition with the MED nodes were tested using Analysis of Similarities (ANOSIM). Interaction between oral malodour and the different cohorts in the subgingival, interdental and tongue microbiota were explored using Permutational Multivariate Analysis of Variance (PERMANOVA). Dissimilarities and distances used in the testing for differences included beta diversity indices such as Bray-Curtis, Horn, Jaccard, Kulczynski and Simpson. The 16S data partitioned by MED were further analysed using the Maximal Information-Based Nonparametric Exploration (MINE) statistic in the R-package minerva [41], to explore associations and interactions between the bacterial species detected in the samples. Pairs that yielded MIC values >0.3 were explored further. Non-parametric correlation analyses were performed using GraphPad Prism 8.0 (GraphPad Software Inc, USA) for Microsoft Windows.

## RESULTS

A total of 110 samples from 51 participants overall were included in the final analysis. The MED pipeline analysed an aggregate of 7,552,373 quality filtered sequences, with the final topology featuring 1,251 nodes that mapped to 255 human oral taxa at ≥98.5% identity.

### Relationship between oral bacterial alpha diversity, clinical indices and VSCs

Subgingival plaque alpha diversity (as measured by the Shannon index) in the study population showed a stronger positive association to hydrogen sulfide in the mouth air (r=0.29), than clinical measurements of disease such as bleeding on probing (BOP; r=0.01) or percentage of periodontal pockets ≥5mm (r=0.15; Figure 1a). Subgingival plaque also showed a positive association with the Plaque Index (r=0.37; p=0.049).

**Figure 1.**
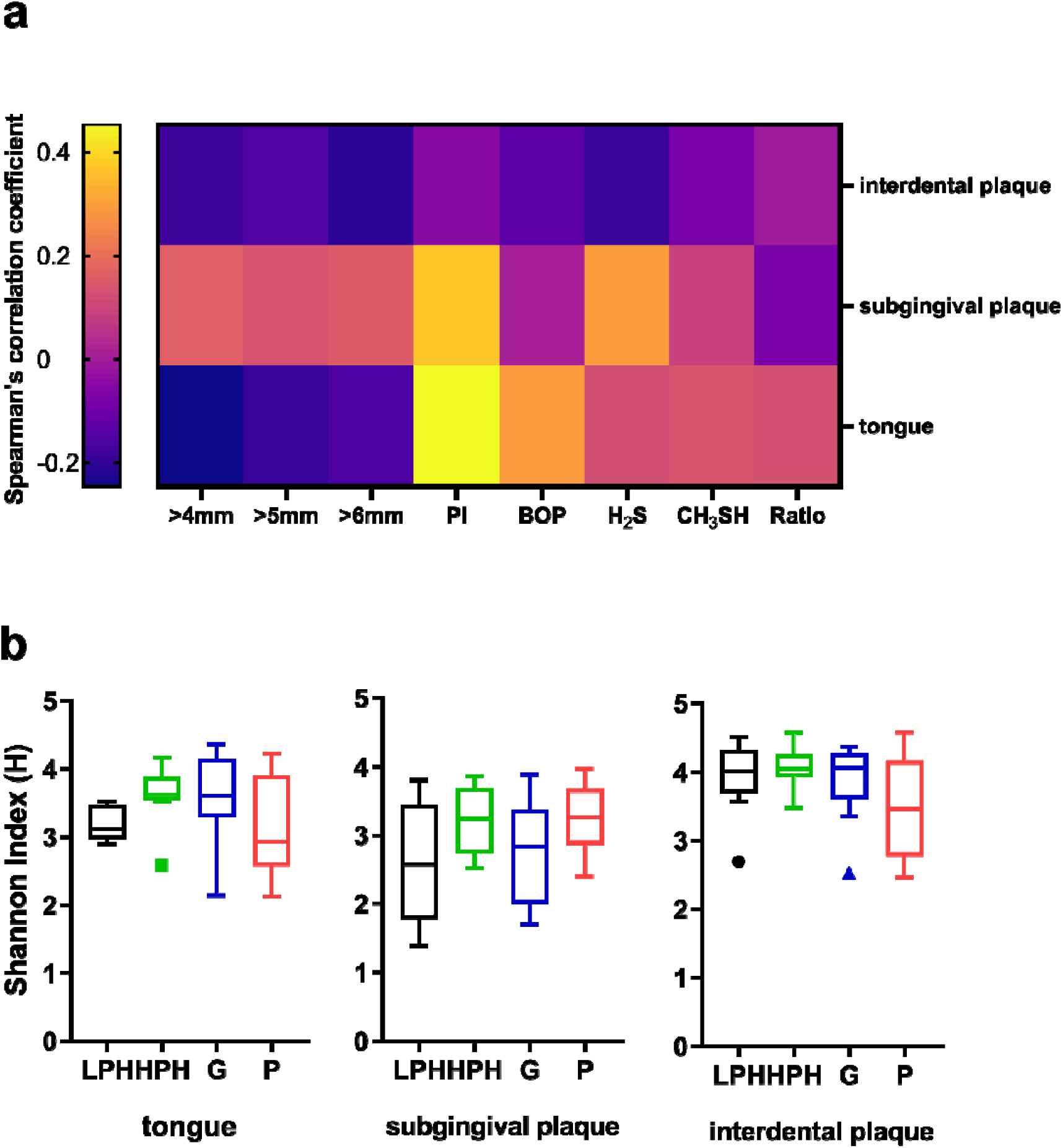
**(a) Heatmap showing the correlation between the Shannon diversity index values of the tongue, subgingival & interdental plaque and the percentage of probing pocket depths ≥4mm, ≥5mm, ≥6mm, plaque index (PI), bleeding on probing (BOP), concentration of H_2_S, CH_3_SH and CH_3_SH: H_2_S. Significance (p-values) of all the correlations are listed in the supplemental files (b) Box plots of Shannon diversity index values from the different disease groups for tongue, subgingival plaque and interdental plaque (LPH=Low plaque health; HPH=High plaque health, G=Gingivitis; P=Periodontitis). Midline denotes median and the boxes extend from the 1**^st^ **and 3**^rd^ **quartile.**

Although statistically not significant, median subgingival and interdental plaque diversities increased and decreased respectively in periodontitis compared to the healthy (Figure 1b), and this relationship was mirrored in their association with the VSCs (Figure 1a). The interdental plaque diversity as measured by the Shannon index showed weak negative associations with most of the clinical and breath measurements (Figure 1a), with the strongest associations to probing depths >6mm (r=-0.21) and the VSC H_2_S (r=-0.19).

Elevated hydrogen sulfide and methanethiol concentrations and an increased methanethiol to hydrogen sulfide ratio (CH_3_SH: H_2_S) were observed in the mouth air of periodontitis patients in comparison to healthy patients (Table 1). Elevated CH_3_SH: H_2_S was statistically significant in the comparison between low plaque health and periodontitis (p=0.04) and not significant in the comparison with gingivitis (p=0.06). All other comparisons were not significant.

### Relationship between oral malodour and the periodontal microbiota

The subgingival and interdental plaque communities were found to be closely related in terms of their shared species (Figure 2b; Jaccard F=26.22, p<0.001) and differed more in the relative abundances of their microbiota (Figure 2a; Bray-Curtis F=16.48, p<0.001). The interdental and subgingival plaque microbial beta diversities were explored using a PERMANOVA model with two factors, namely, presence of oral malodour and the clinical diagnosis, to ascertain their relative contribution in explaining the underlying variation in the samples (Figure2c-f). Clinical diagnosis was observed to be more significant than oral malodour for both interdental and subgingival plaque, and no statistically significant interaction between the two factors was detected. A significant difference was detected between the cohorts among the interdental plaque samples (F=1.07, p=0.05; Figure 2c), with the presence or absence of oral malodour not significant (F=1.13, p=0.1; Figure 2d). Similarly, clinical diagnosis was the dominant factor in explaining the variation observed in the subgingival plaque (F=1.00, p=0.01; Figure 2e), with the presence/absence of oral malodour not significant (F=0.71, p=0.3; Figure 2f).

**Figure 2.**
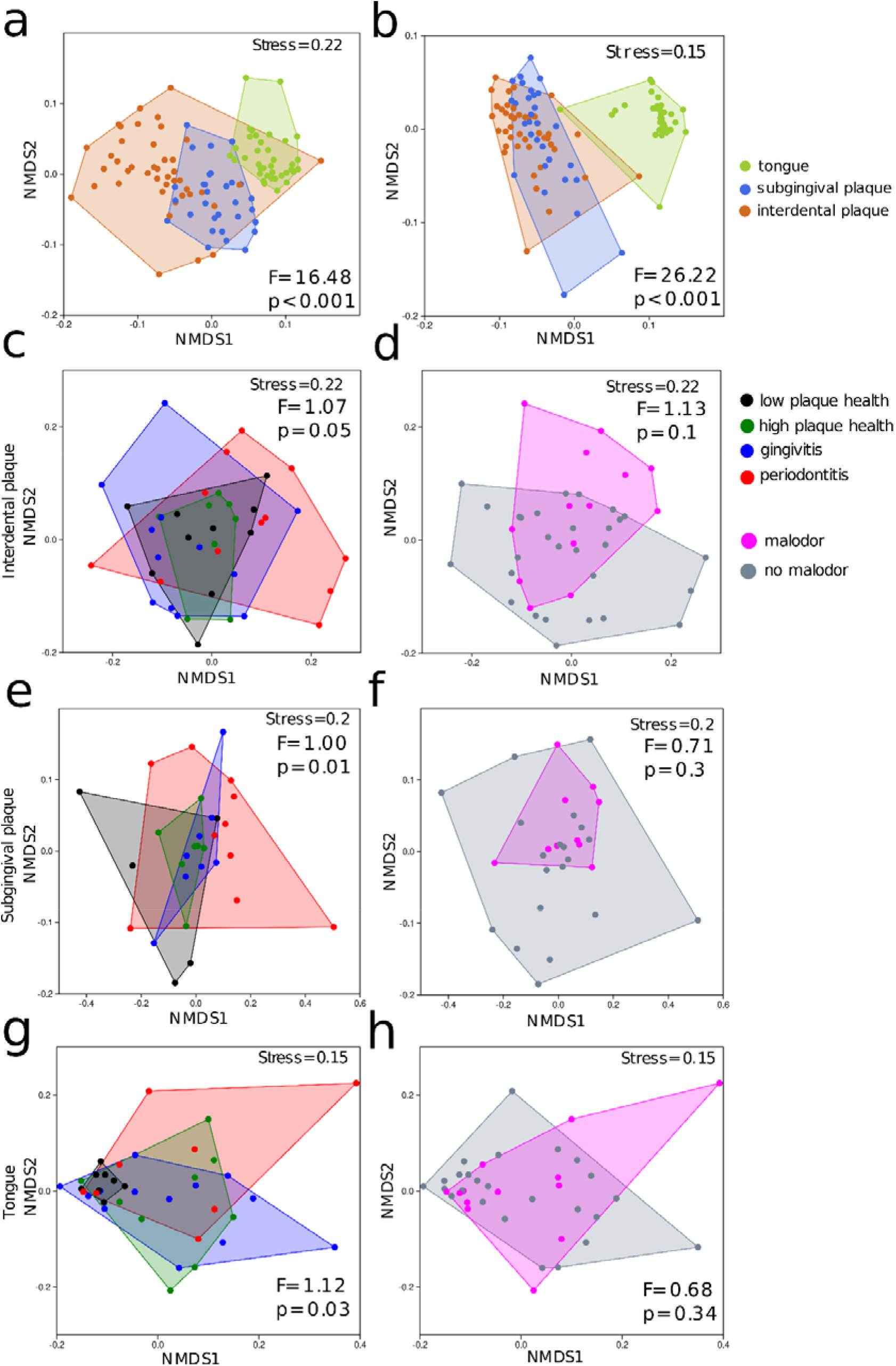
**Non-metric multidimensional scaling (NMDS) plots of the microbiota of tongue, subgingival and interdental plaque based on the Bray-Curtis dissimilarity (a) and Jaccard distance (b), showing significant differences between the sampling sites. NMDS plots of interdental plaque communities based on the Bray-Curtis dissimilarity analysed for differences between the cohorts (c) and presence of oral malodour (d). NMDS plots of subgingival communities based on the Kulczynski distance showing significant differences between the cohorts (e) and oral malodour (f). NMDS plots of tongue communities based on the Bray-Curtis dissimilarity, between the cohorts (g) and oral malodour (h). Convex hulls on all plots contain all data within each grouping.**

Between the periodontal niches, the interdental plaque microbiota differed more between individuals with and without oral malodour than the subgingival plaque in the study population. Microbial species such as *Rothia dentocariosa, Lautropia mirabilis, Leptotrichia hongkongensis, Corynebacterium matruchotii* and *Capnocytophaga* spp decreased in abundance in the interdental plaque of individuals with oral malodour, consistent with the MINE analysis in relation to VSCs (Figure 3). Species such as *Veillonella atypica, Veillonella dispar* and *L. hongkongensis* were present in higher abundances on the tongue of individuals without oral malodour compared to oral malodour in the study population, whereas the most microbial community differences in the subgingival plaque of individuals with oral malodour were due to increased abundance of *Gemella morbillorum, Treponema denticola* and *Peptostreptococcus stomatis* (Figure 3). These microbial species associations were stronger in gingivitis and periodontitis patients with oral malodour, than the healthy.

**Figure 3.**
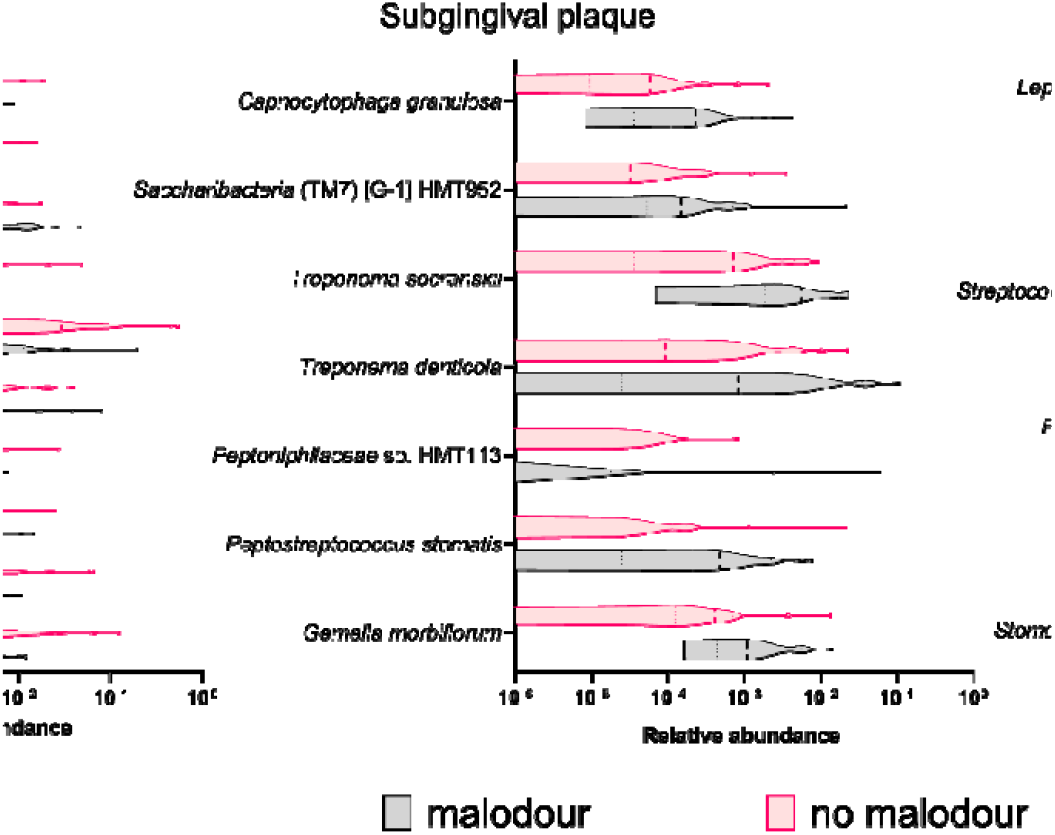
**Violin plots showing relative abundances of taxa that showed associations (MIC >0.3) with VSCs in the MINE analysis in the different niches, with the samples assigned to malodour or no malodour based on the detection threshold as stated in the Methods.**

#### Relationship between the tongue microbiota and periodontitis

The tongue had the lowest species richness (mean = 115 ± 20) in comparison to subgingival (mean = 151 ± 23) and interdental plaque (mean = 162 ± 18). Multivariate ordination showed that the tongue was a more distinct niche in terms of its overall community structure compared to the subgingival and interdental niches (Figure 2a Bray-Curtis F=16.48, p<0.001 & 2b, Jaccard F=26.22, p<0.001). The tongue also showed significant changes in its community structure in relation to health, gingivitis and periodontitis than oral malodour (F=1.12, p=0.03; Figure 2g & 2h). However, no significant interaction between malodour and the clinical diagnosis was detected from the tongue samples.

### Microbial species associated with clinical indices of periodontitis

Dental plaque coverage and BOP showed significant associations to several microbial taxa on the tongue dorsum from the MINE analysis (the full list of taxa is shown in the Supplemental file). *Rothia mucilaginosa, Gemella sanguinis, Peptostreptococcaceae* [XI][G-1] sulci and Lachnospiraceae [G-2] HMT 096, were associated with both plaque coverage and BOP. *Actinomyces* HMT 180, *Leptotrichia* spp., *Prevotella* HMT 313, *Saccharibacteria* (TM7) [G-1] HMT 352, *Peptostreptococcaceae* [XI][G-1] sulci, *Haemophilus parainfluenzae* and *Atopobium parvulum* were associated with dental plaque coverage, whereas species such as Streptococcus HMT 074, *Actinomyces* HMT 172, *Fusobacterium periodonticum, Veillonella atypica* and *Solobacterium moorei* were associated with an increase in BOP. The detection of R. *mucilaginosa* sequence variants by MED in the different niches showed that the proportions of the different variants in the tongue was more closely related to the subgingival niche (ANOSIM-Bray-Curtis r=0.17, p<0.001), than the interdental niche (Figure 4; ANOSIM-Bray-Curtis r=0.76, p<0.001).

**Figure 4.**
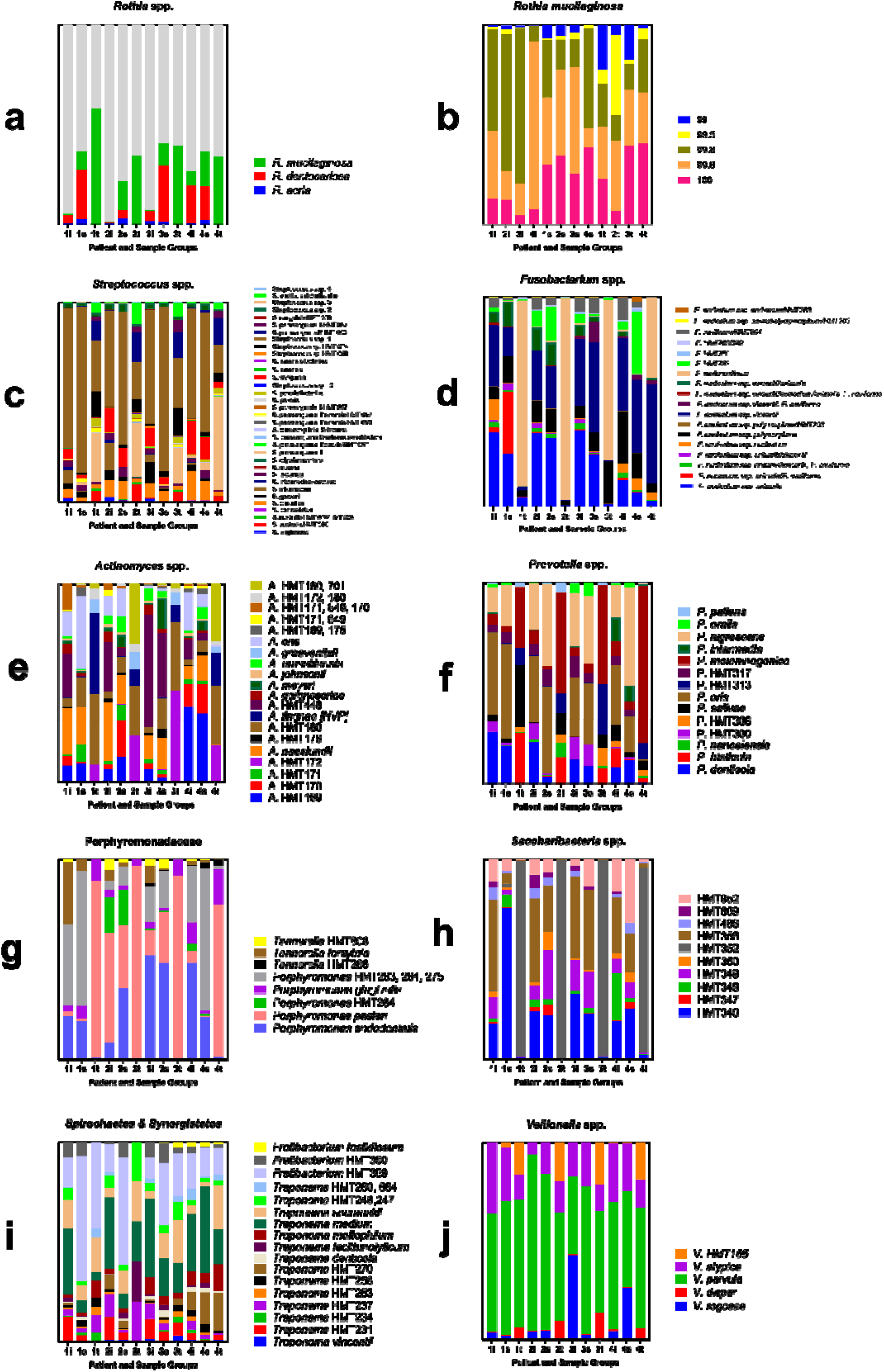
**Relative abundance of Rothia spp. (a), sequence variant diversity of *Rothia mucilaginosa* (legend indicates sequence identity match to strain DY-18) normalised to its total sequence variant abundance (b), Abundance changes of each microbial species normalized to total abundance within the genus, family or phyla: *Streptococcus* spp. (c), *Fusobacterium* spp. (D), *Actinomyces* spp. (E), *Prevotella* spp. (f), Porphyromonadaceae (g), *Saccharibacteria* spp. (h), Spirochaetes & Synergistetes (i) and *Veillonella* spp. (j) between low plaque health (1), high plaque health (2), gingivitis (3) and periodontitis (4) in tongue (t), interdental (i) and subgingival plaque (s). Unique nodes that matched to several different members of *Streptococcus* spp are denoted by numbers in the legend.**

Among the measured clinical indices, the strongest association was observed between subgingival plaque diversity and the percentage of dental plaque coverage (Figure 1a), with a number of *Streptococcus* spp. involved in the differences observed, including *S. sanguinis, S. parasanguinis II, S. cristatus* (clade 578), *S. lactarius* and *S. anginosus* (Figure 4). Neisseria *flavescens, N. subflava, N. elongata, Rothia dentocariosa, Leptotrichia wadei, L. hongkongensis, Bacteroidales* [G-2] HMT 274, *Gemella morbillorum, Granulicatella adiacens* and *Prevotella oris* are other species that showed important shifts in the subgingival plaque in association with increasing plaque coverage (Supplemental files).

The decrease in the interdental alpha diversity in association with an increase in periodontal pockets (≥6mm) was represented by phylotypes such as *Tannerella* HMT 808, *Lautropia mirabilis, Saccharibacteria* HMT 349 & HMT 869, *Fusobacterium* spp., *P. nigrescens, P. oris, P. endodontalis* and *S. moorei* (Figure 4). A list of all associations between bacterial taxa and clinical and breath measurements detected by MINE analysis with the MIC value >0.3, and all multiple comparisons within genera in Figure 4 that were significant are listed in the Supplemental files.

## DISCUSSION

The role of the tongue biofilm in oral malodour has been well established, with the presence of VSCs such as hydrogen sulfide and methanethiol being key markers of oral malodour [10, 18]. The findings of the present study highlight potentially important associations between microbial diversity in the periodontal environment and malodour exhibited in individuals with periodontal health, gingivitis and periodontitis (Figure 1 & Figure 2). Consistent with the literature, elevated concentrations of the VSC methanethiol was observed in the chronic periodontitis and gingivitis cohorts compared to health [1,3]. Clinically healthy individuals with high plaque coverage also had an elevated CH_3_SH: H_2_S ratio in the present study, suggesting possible increased disease activity in these individuals. The positive association between the subgingival plaque diversity and the VSC H_2_S (Figure 1) suggests that oral malodour may be a sign of a dysbiotic oral microbiota, given that higher subgingival plaque diversity is associated with dysbiosis and periodontitis [7].

Two previous studies that investigated the tongue microbiota in relation to oral malodour using NGS did not find an association with microbial diversity on the tongue [13,14]. One study relied on a H_2_S concentration threshold to detect absence of malodour and moderate/severe malodour similar to the present study, whereas the more recent study of the tongue microbiota used organoleptic assessments to detect oral malodour [13, 14]. Consistent with both studies, the present study also did not find any significant association between VSCs and the alpha diversity in the tongue (Figure 1). In addition, the tongue microbial species observed to vary in abundance between individuals with and without oral malodour in the present study (Figure 3), are broadly consistent with the study that used organoleptic measures to recruit individuals with oral malodour [14]. This emphasizes the strong association between an organoleptic assessment of oral malodour and VSCs in the oral cavity.

The general prevalence rates of malodour ranged between 27-35% in the current study cohorts (Table 1). Given the exclusion criteria of this study, this is not in contrast to the literature where prevalence of periodontitis in conjunction with oral malodour is not reported to be high [19, 20]. However, individuals with gingivitis or periodontitis with the presence of VSCs were measured at a higher concentration than the healthy in the present study. Further, clinical indices such as plaque coverage and BOP were found to be associated with tongue alpha diversity (Figure 1a).

Data from the present study suggested that the tongue was much more distinct in its bacterial community structure than the subgingival or interdental plaque, which is consistent with the literature (Figure 2a & 2b) [21]. However, it was found that bacteria normally resident in the periodontal niches, increased in relative abundance on the tongue with higher plaque coverage and BOP values in the study population (Figure 1 & Figure 4). This suggests that the tongue dorsum may become a reservoir for periodontal bacteria possibly due to factors such as better nutrient availability resulting from periodontal inflammation and changes in redox potential could form part of the ecological pressures that could allow the tongue to support a more diverse and periodontopathic microbial population [42].In turn, the tongue may also play a role in helping these microbial species in recolonising the subgingival and interdental niches.

It has been previously reported that interdental ‘floss odor’ had a small but significant contribution to VSC concentration in the oral cavity and overall oral malodour [19]. In our analysis of the interdental and subgingival microbiota, the presence of oral malodour explained more of the existing variation in the interdental plaque community structure than the subgingival plaque (Figure 2 c – 2f). The microbial community observed in the interdental plaque was more diverse than subgingival plaque or the tongue (Figure 1b) and harboured a relatively higher abundance of known VSC producing bacteria in the current study population, suggesting a potential link between interdental plaque and oral malodour [22].

The tongue may be the dominant niche responsible for malodour, however, the associations found in the present study suggest that subgingival microbiota may have a role to play in influencing the composition of the tongue microbiota. For instance, MINE analysis suggested that the nitrate reducing *R. mucilaginosa*, present in high relative abundance and prevalence in the tongue of individuals with periodontal health and low plaque (<30% plaque coverage) showed a decrease in healthy individuals that exceeded this plaque coverage threshold (Figure 4) [23]. *Rothia* spp are generally health associated, and it is possible that tongue dwelling *R. mucilaginosa* is particularly sensitive to subclinical inflammation resulting from dental plaque accumulation. This is consistent with results from a recent study that showed that *Rothia* spp in subgingival plaque are significantly inhibited at the onset of experimental gingivitis induced in humans [43]. MED analysis found five major sequence variants of *R. mucilaginosa* in all the samples, with the composition in the tongue and subgingival niches much more closely related across the different disease cohorts, than the interdental plaque, further indicating a possible temporal relationship between the tongue dorsum and subgingival plaque (Figure 4).

Similarly, the interdental plaque had the highest abundance and diversity of fusobacteria among the niches studied, with the tongue dorsum harbouring an uneven community of the same genus, with *F. periodonticum* being the dominant species (Figure 4). In gingivitis and periodontitis, a major shift was observed in the community composition, with an increase of *F. nucleatum* ssp *polymorphum* and *F. nucleatum* ssp vincentii, respectively on the tongue reflecting the role of interdental plaque in influencing both the subgingival and tongue microbiota in disease. In particular, the presence of *F. nucleatum* ssp *polymorphum* and *F. nucleatum* ssp *vincentii* in high abundances in the interdental and subgingival plaque but not in the tongue of healthy individuals with low plaque scores suggests a significant change in the aforementioned factors such as nutrient availability and redox potential on the tongue in periodontitis resulting in a more diverse ecology. Furthermore, *S. sanguinis* was present in all cohorts at a higher abundance in the interdental plaque but was observed to increase with disease in the subgingival plaque and tongue, suggesting the route of colonisation in disease via the interdental plaque (Figure 4). Conversely, *S. parasanguinis* II was observed in high abundance in the tongue, with the bacterium increasing in abundance in the subgingival and interdental plaque in gingivitis and periodontitis (Figure 4). The species *Porphyromonas pasteri* (HMT 279), also showed a striking increase in abundance in the interdental and subgingival niches of healthy individuals with high plaque, gingivitis and periodontitis compared to low plaque health, where it was normally prevalent only on the tongue (Figure 4), highlighting the population dynamics that can occur in healthy individuals with high plaque index. *Treponema lecithinolyticum* and T. medium increased in abundance in the tongue of high plaque healthy but were normally prevalent in the subgingival and interdental niches. It is possible that these different community composition differences between the different genera are related, as the role of streptococci in aggregating with fusobacteria has been documented previously [24,25]. These data may also point to specific roles of these microbial species in different disease states, as not all species were observed to be present in a higher abundance in a different niche in the different disease cohorts compared to health. For example, species belonging to Prevotella spp., Veillonella spp., Saccharibacteria (TM7) spp., Actinomyces spp., and betaproteobacteria such as Neisseria spp, Kingella oralis, Lautropia mirabilis and Eikenella corrodens, showed niche specificity irrespective of disease state, and did not appear to crossover to a different niche in disease (Figure 4; supplement file). These observations may also explain the observed differences in the VSCs and the CH_3_SH: H_2_S, where malodour in healthy individuals was predominantly due to H_2_S, with CH_3_SH being more prevalent in the disease groups.

Whilst the sample size for this study is comparable to other reported oral malodour studies [13, 14], the study has small numbers of participants within the cohorts when stratified into presence or absence of malodour (Figure 3 & Supplementary Figure S2). This could limit the significance of different microbial species that can be detected statistically to be different within the cohorts. Nevertheless, a model for the role of oral microbiota in oral malodour may be synthesised from observations made in this study and other studies in the literature [13-15, 22, 26-28] (Figure 5), whereby the tongue and periodontal microbiota are subject to bidirectional transmission of species that alter the ecology of both niches in association with increased indicators of periodontitis such as plaque accumulation, bleeding on probing and increased periodontal probing depths. This altered ecology and habitat play a role in contributing to oral malodour in disease. However, in health, the oral malodour is caused by a different oral ecological landscape, predominantly driven by the tongue microbiota.

**Figure 5.**
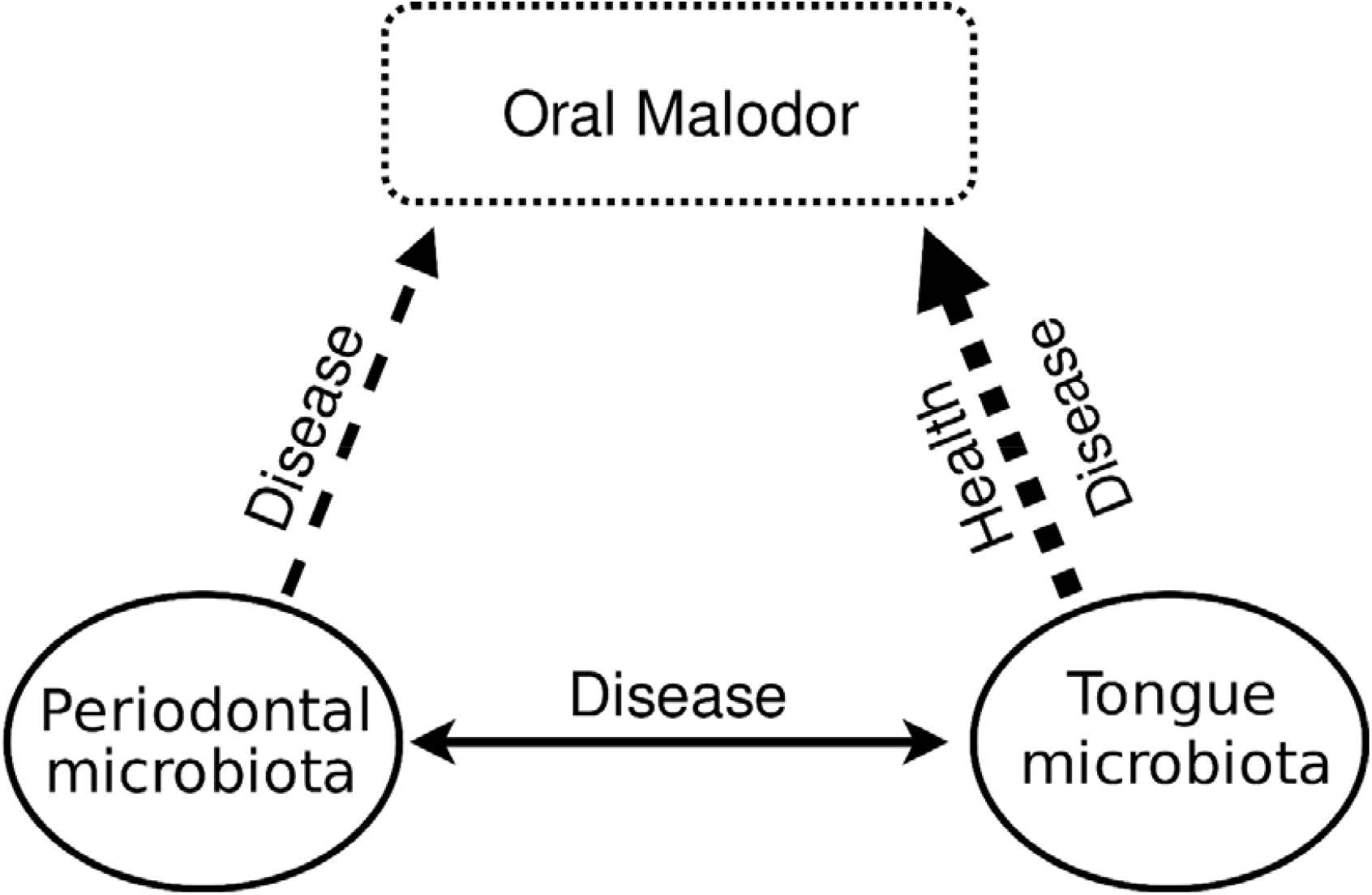
**A conceptual model of pathological oral malodour as suggested by the oral microbial ecological shifts observed in health, gingivitis and chronic periodontitis.**

In conclusion, the periodontal microbiota was associated with oral malodour in health and disease, with the tongue microbial community structure shifting significantly in association with clinical signs of periodontitis. The increased presence of periodontopathic bacteria on the tongue dorsum of individuals with periodontitis, partly explains the increased VSC concentrations in the mouth air of individuals with periodontitis. These data further suggest that it may be important to reduce dental plaque accumulation to combat oral malodour. In addition, the tongue may play a key role in periodontitis by supporting colonisation of periodontopathic bacteria. Therefore, it may be useful to determine in further studies if treatment of periodontitis should also include measures to reduce the tongue bacterial load.

## Data Availability

Raw sequences analysed in this study are available in the NCBI Sequence Read Archive accession: PRJNA649803

