## Supplemental file for "Interdental and subgingival microbiota may affect the tongue microbial ecology and oral malodour in health, gingivitis and periodontitis"

**Supplementary Figure S1. Flow chart of the clinical protocol sequence with the oral examination and the different sampling aspects.**

Informed Consent

Participant information sheet and consent forms

Screening & Enrolment

Medical history questionnaire

Last oral activity

Demographics questionnaire

Sample collection

Breath sample and analysis

Saliva

Tongue biofilm

Oral examination

Full mouth six point pocket charting

Determination of diagnosis

one tooth per quadrant chosen for plaque collection

Sample collection

Supragingival plaque

Subgingival plaque

interdental plaque

Oral examination

Full mouth Plaque Index measurement

Participant debrief

End of clinical protocol

**Supplementary Figure S2. Comparison of Shannon diversity index values for the tongue, subgingival and interdental plaque of individuals with or without oral malodour within the different cohorts: Low plaque health (LPH), High plaque health (HPH), Gingivitis (G) and Periodontitis (P). Median and range values are shown.**

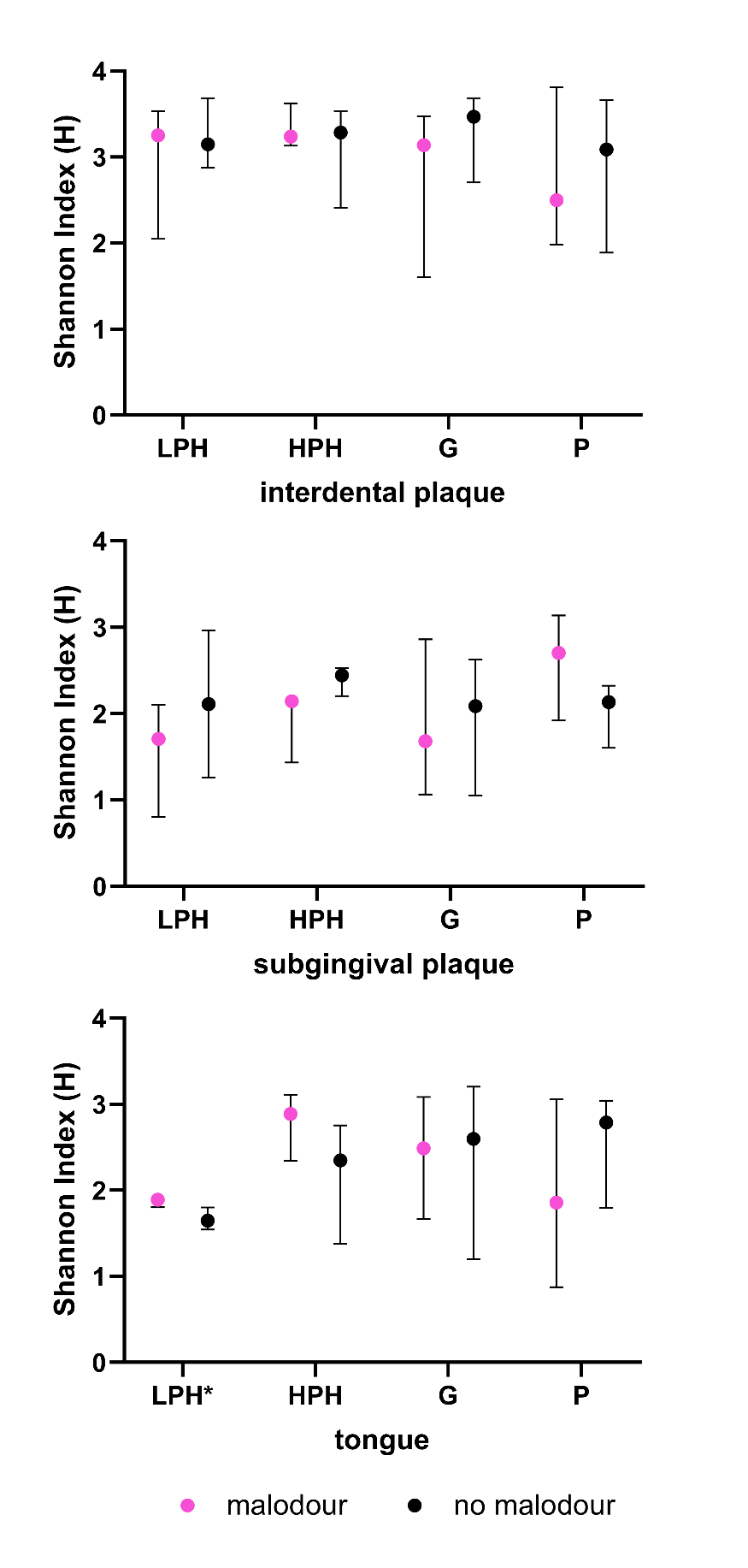

**Supplementary Figure S3. Box plots of Shannon diversity index values calculated from samples rarefied to the lowest library size (14000 reads) (LPH=Low plaque health; HPH=High plaque health, G=Gingivitis; P=Periodontitis). Midline denotes median and the boxes extend from the 1st and 3rd quartile.**

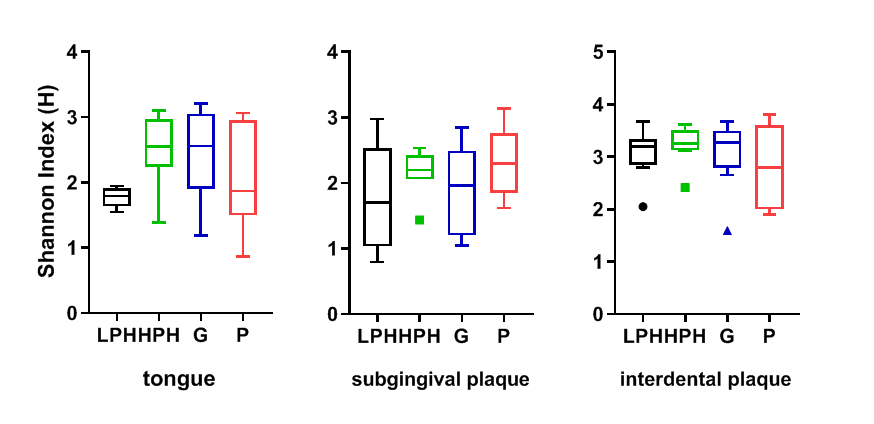

**Supplementary Figure S4. Box plots of Chao1 values for the tongue, subgingival and interdental plaque of the Low plaque health (LPH), High plaque health (HPH), Gingivitis (G) and Periodontitis (P) cohorts. Midline denotes median and the boxes extend from the 1st and 3rd quartile.**

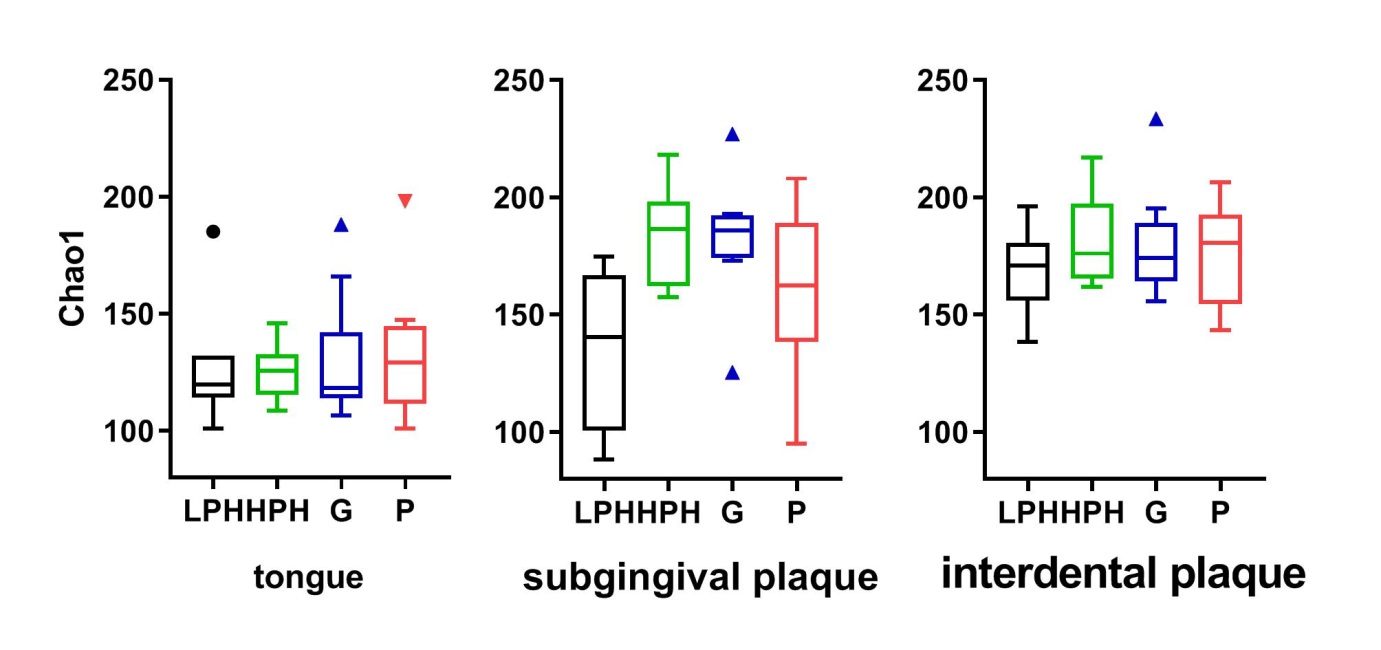

**Supplementary Figure S5. Box plots of Simpson index values for** **the tongue, subgingival and interdental plaque of the Low plaque health (LPH), High plaque health (HPH), Gingivitis (G) and Periodontitis (P) cohorts. Midline denotes median and the boxes extend from the 1st and 3rd quartile.**

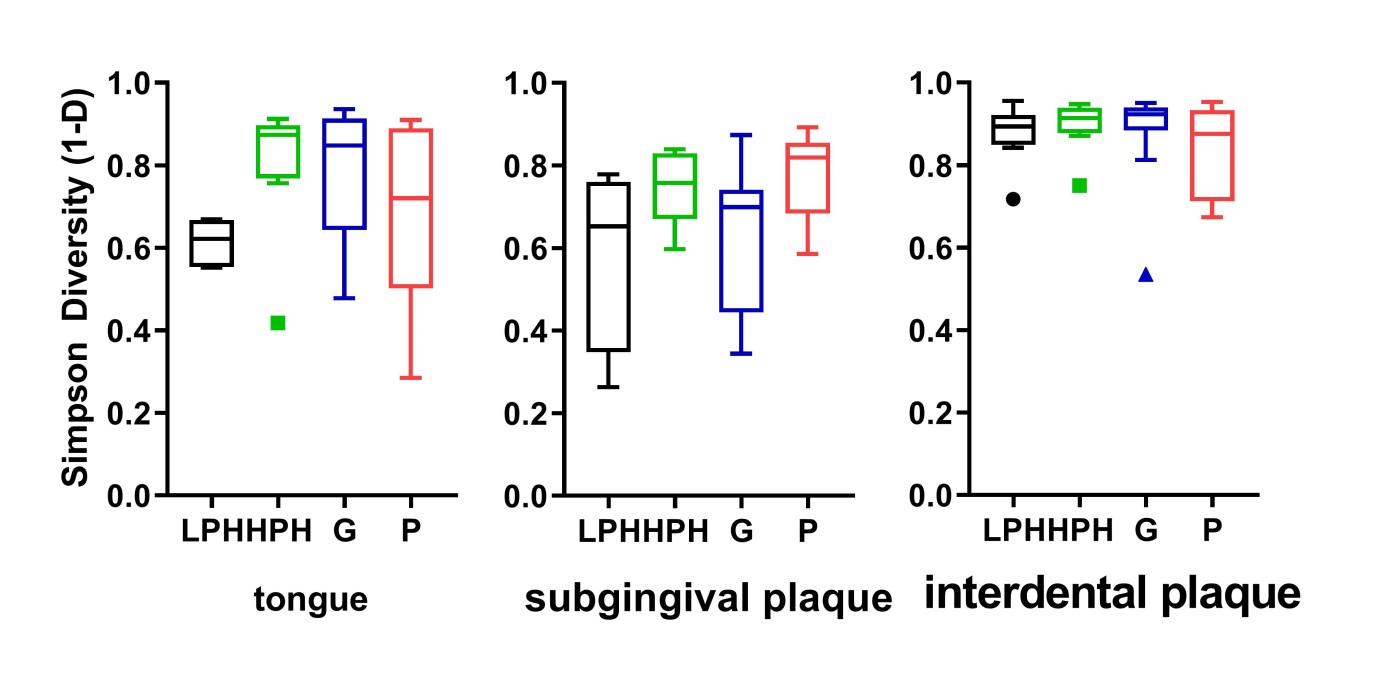

**Supplementary Figure S6. Box plots of Shannon diversity index values for the tongue, subgingival and interdental plaque in the study population using the 2017 classification for health (H), gingivitis (G) and periodontitis (P). Midline denotes median and boxes extend from the 1^st^ and 3^rd^ quartile.**

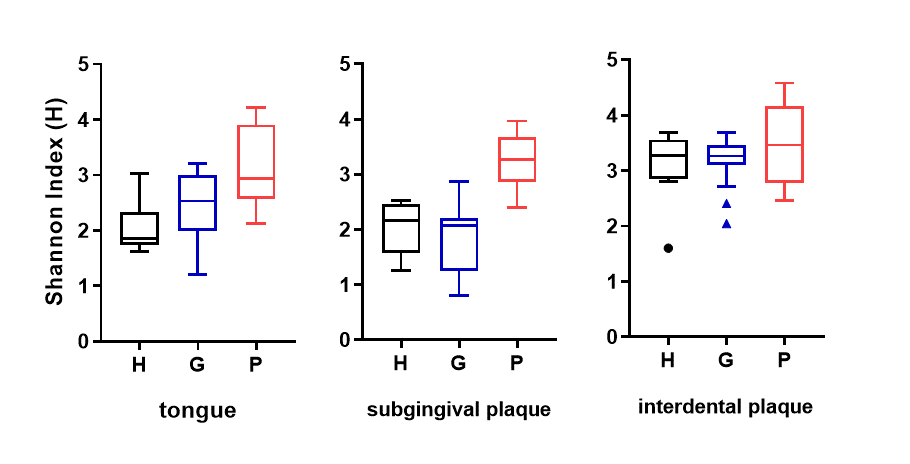

**Supplementary Table S1. Table listing uncorrected p-values for the comparisons shown in Figure 1a.**

|  | Interdental plaque | | Subgingival plaque | | Tongue | |
| --- | --- | --- | --- | --- | --- | --- |
| **Clinical & breath variables** | **Spearman r**  **(95% CI)** | **P-value**  **(two-tailed)** | **Spearman r**  **(95% CI)** | **P-value**  **(two-tailed)** | **Spearman r**  **(95% CI)** | **P-value**  **(two-tailed)** |
| **>4mm** | **-0.182**  **(-0.467 to 0.138)** | **0.249** | **0.161**  **(-0.222 to 0.501)** | **0.396** | **-0.245**  **(-0.531 to 0.0903)** | **0.138** |
| **>5mm** | **-0.152**  **(-0.443 to 0.169)** | **0.337** | **0.130**  **(-0.252 to 0.477)** | **0.492** | **-0.192**  **(-0.49 to 0.145)** | **0.248** |
| **>6mm** | **-0.209**  **(-0.489 to 0.11** | **0.184** | **0.155**  **(-0.229 to 0.496)** | **0.415** | **-0.160**  **(-0.464 to 0.178)** | **0.338** |
| **PI** | **-0.050**  **(-0.361 to 0.27)** | **0.754** | **0.368**  **(-0.00973 to 0.654)** | **0.05** | **0.453**  **(0.142 to 0.683)** | **0.005** |
| **BOP** | **-0.134**  **(-0.432 to 0.191)** | **0.405** | **0.008**  **(-0.363 to 0.377)** | **0.967** | **0.294**  **(-0.0385 to 0.567)** | **0.074** |
| **H_2_S** | **-0.191**  **(-0.475 to 0.129)** | **0.225** | **0.291**  **(-0.0884 to 0.597)** | **0.119** | **0.116**  **(-0.221 to 0.428)** | **0.489** |
| **CH_3_SH** | **-0.079**  **(-0.382 to 0.24)** | **0.620** | **0.088**  **(-0.291 to 0.444)** | **0.642** | **0.137**  **(-0.201 to 0.445)** | **0.413** |
| **Ratio** | **-0.005**  **(-0.317 to 0.307)** | **0.974** | **-0.077**  **(-0.435 to 0.302)** | **0.686** | **0.127**  **(-0.21 to 0.437)** | **0.448** |

**Supplementary Table S2. Table listing microbial taxa detected in the interdental plaque that showed the largest loadings across the first two components in a Principal Component Analysis.**

| HMT | Genus | Species |
| --- | --- | --- |
| HMT111 | **Parvimonas** | **micra** |
| HMT169 | **Actinomyces** | **sp._HMT_169** |
| HMT200 | **Fusobacterium** | **nucleatum ssp vincentii** |
| HMT202 | **Fusobacterium** | **nucleatum ssp polymorphum** |
| HMT204, 689 | **Fusobacterium** |  |
| HMT213 | **Leptotrichia** | **hongkongensis** |
| HMT346 | **Saccharibacteria (TM7) [G-1]** | **HMT 346** |
| HMT356 | **Saccharibacteria (TM7) [G-5]** | **HMT 356** |
| HMT420 | **Fusobacterium** | **nucleatum ssp animalis** |
| HMT476 | **Neisseria** | **subflava** |
| HMT512, 762 | **Aggregatibacter** | **segnis/sp._HMT_512** |
| HMT578 | **Streptococcus** | **cristatus clade578** |
| HMT584 | **Treponema** | **denticola** |
| HMT587 | **Rothia** | **dentocariosa** |
| HMT595 | **Corynebacterium** | **durum** |
| HMT598 | **Neisseria** | **elongata** |
| HMT681 | **Rothia** | **mucilaginosa** |
| HMT718 | **Haemophilus** | **parainfluenzae** |
| HMT758 | **Streptococcus** | **sanguinis** |
| HMT893 | **Actinomyces** | **oris** |

**Supplementary Table S3. Table listing microbial taxa detected in the subgingival plaque that showed the largest loadings across the first two components in a Principal Component Analysis.**

| HMT | Genus | Species |
| --- | --- | --- |
| HMT070, 398, 058, 677, 707, 064, 638, 071, 061, 431, 423 | **Streptococcus** | |
| HMT110, 393 | **Parvimonas** | |
| HMT112 | **Peptostreptococcus** | **stomatis** |
| HMT188 | **Rothia** | **aeria** |
| HMT411 | **Streptococcus** | **parasanguinis_clade_411** |
| HMT420, 689 | **Fusobacterium** | |
| HMT534 | **Granulicatella** | **adiacens** |
| HMT578 | **Streptococcus** | **cristatus clade 578** |
| HMT587 | **Rothia** | **dentocariosa** |
| HMT587, 188 | **Rothia** |  |
| HMT681 | **Rothia** | **mucilaginosa** |
| HMT718 | **Haemophilus** | **parainfluenzae** |
| HMT74 | **Streptococcus** | **sp. HMT 074** |
| HMT758 | **Streptococcus** | **sanguinis** |

**Supplementary Table S4. Table listing microbial taxa detected in the tongue biofilm samples that showed the largest loadings across the first two components in a Principal Component Analysis.**

| HMT | Genus | Species |
| --- | --- | --- |
| HMT021, 755 | **Streptococcus** | |
| HMT066, 721 | **Streptococcus** | |
| HMT070, 398, 058, 677, 707, 064, 638, 071, 061, 431, 423 | **Streptococcus** | |
| HMT071, 398, 058, 707, 728, 070, 423, 677, 064, 061, 638, 431 | **Streptococcus** | |
| HMT172 | **Actinomyces** | **sp. HMT 172** |
| HMT180 | **Actinomyces** | **sp. HMT 180** |
| HMT313 | **Prevotella** | **sp. HMT 313** |
| HMT411 | **Streptococcus** | **parasanguinis clade 411** |
| HMT411, 767, 057 | **Streptococcus** | |
| HMT417 | **Leptotrichia** | **sp. HMT 417** |
| HMT469 | **Prevotella** | **melaninogenica** |
| HMT476 | **Neisseria** | **subflava** |
| HMT534 | **Granulicatella** | **adiacens** |
| HMT681 | **Rothia** | **mucilaginosa** |
| HMT718 | **Haemophilus** | **parainfluenzae** |
| HMT723 | **Atopobium** | **parvulum** |
| HMT74 | **Streptococcus** | **sp. HMT 074** |
| HMT948 | **Streptococcus** | **lactarius** |

**Supplementary Table S5. Tables listing F values of pairwise PERMANOVA analysis of the community structure in the different niches. *Bonferroni-corrected p<0.05**

| **Tongue**  **Bray Curtis (Jaccard)** | **Periodontitis** | **Low plaque health** | **Gingivitis** | **High plaque health** |
| --- | --- | --- | --- | --- |
| **Periodontitis** |  | **2.632 (1.724)** | **0.7865 (1.417)** | **0.6261 (1.836*)** |
| **Low plaque health** | **2.632 (1.724)** |  | **3.231 (1.57)** | **4.611* (1.242)** |
| **Gingivitis** | **0.7865 (1.417)** | **3.231 (1.57)** |  | **0.6492 (0.9021)** |
| **High plaque health** | **0.6261 (1.836*)** | **4.611* (1.242)** | **0.6492 (0.9021)** |  |

| **Subgingival plaque**  **Bray Curtis (Jaccard)** | **Periodontitis** | **Low plaque health** | **Gingivitis** | **High plaque health** |
| --- | --- | --- | --- | --- |
| **Periodontitis** |  | **1.276 (1.775)** | **1.004 (1.336)** | **1.706 (1.471)** |
| **Low plaque health** | **1.276 (1.775)** |  | **0.6447 (1.915)** | **1.791 (2.109*)** |
| **Gingivitis** | **1.004 (1.336)** | **0.6447 (1.915)** |  | **0.8517 (0.7204)** |
| **High plaque health** | **1.706 (1.471)** | **1.791 (2.109*)** | **0.8517 (0.7204)** |  |

| **Interdental plaque**  **Bray Curtis (Jaccard)** | **Periodontitis** | **Low plaque health** | **Gingivitis** | **High plaque health** |
| --- | --- | --- | --- | --- |
| **Periodontitis** |  | **1.469 (1.847)** | **1.661 (1.207)** | **1.743 (1.553)** |
| **Low plaque health** | **1.469 (1.847)** |  | **1.113 (1.374)** | **0.7393 (1.286)** |
| **Gingivitis** | **1.661 (1.207)** | **1.113 (1.374)** |  | **1.251 (0.9834)** |
| **High plaque health** | **1.743 (1.553)** | **0.7393 (1.286)** | **1.251 (0.9834)** |  |

**Supplementary Table S6. Table listing all correlations detected by MINE analysis between clinical/breath measurements and bacterial taxa in the tongue.**

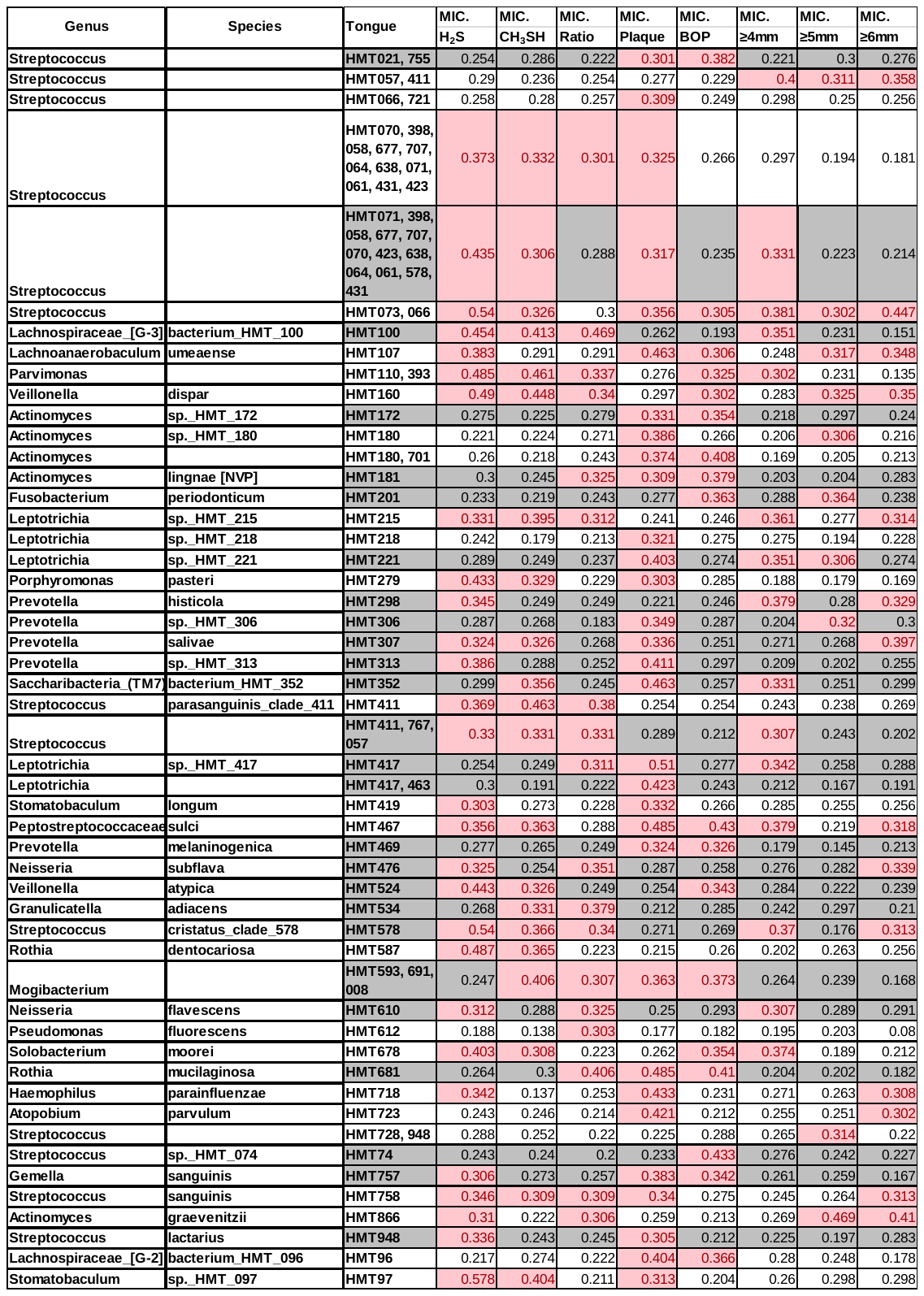

**Supplementary Table S7. Table listing all correlations detected by MINE analysis between clinical/breath measurements and bacterial taxa in the subgingival plaque.**

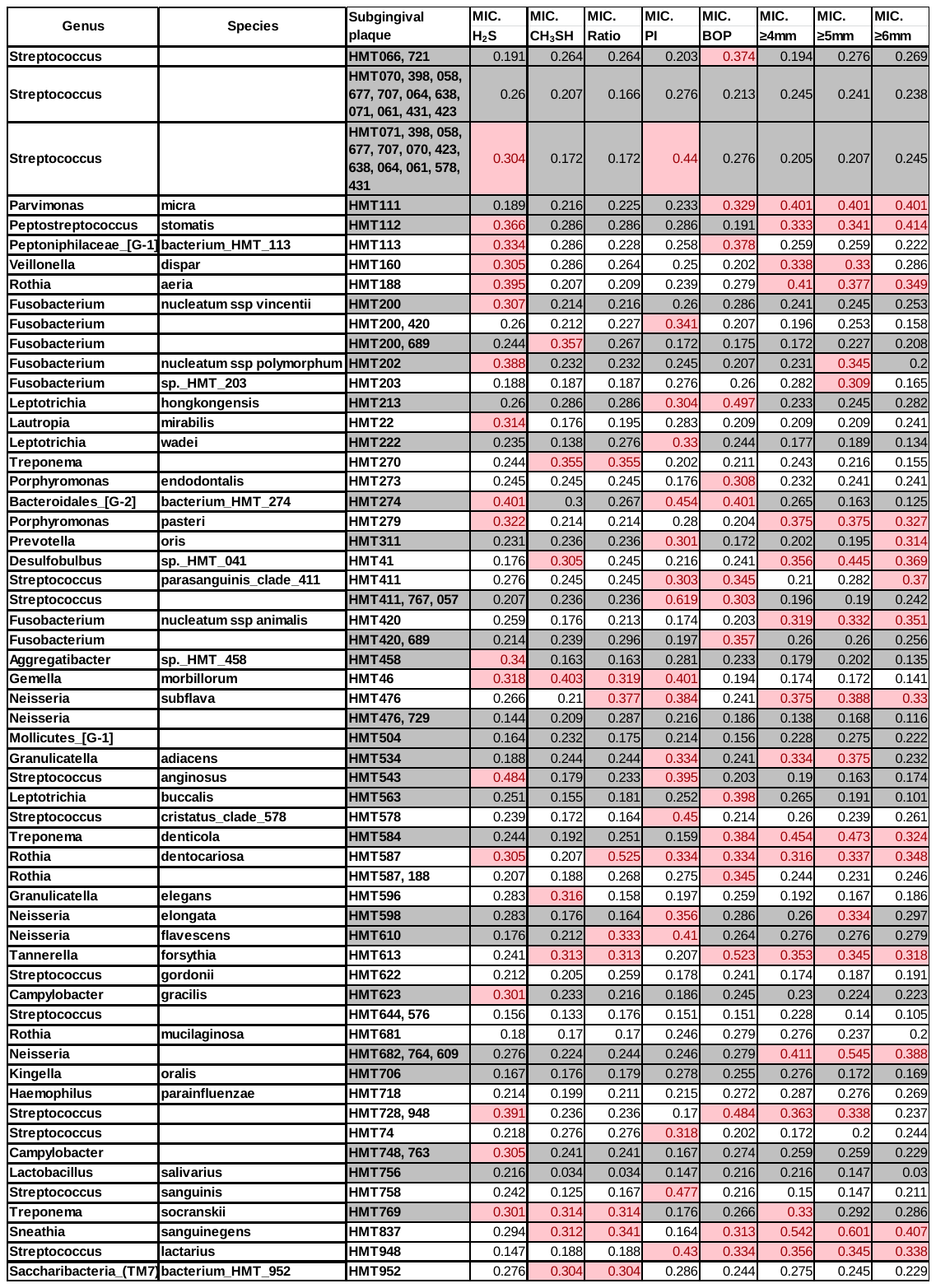

**Supplementary Table S8. Table listing all correlations detected by MINE analysis between clinical/breath measurements and bacterial taxa in the interdental plaque.**

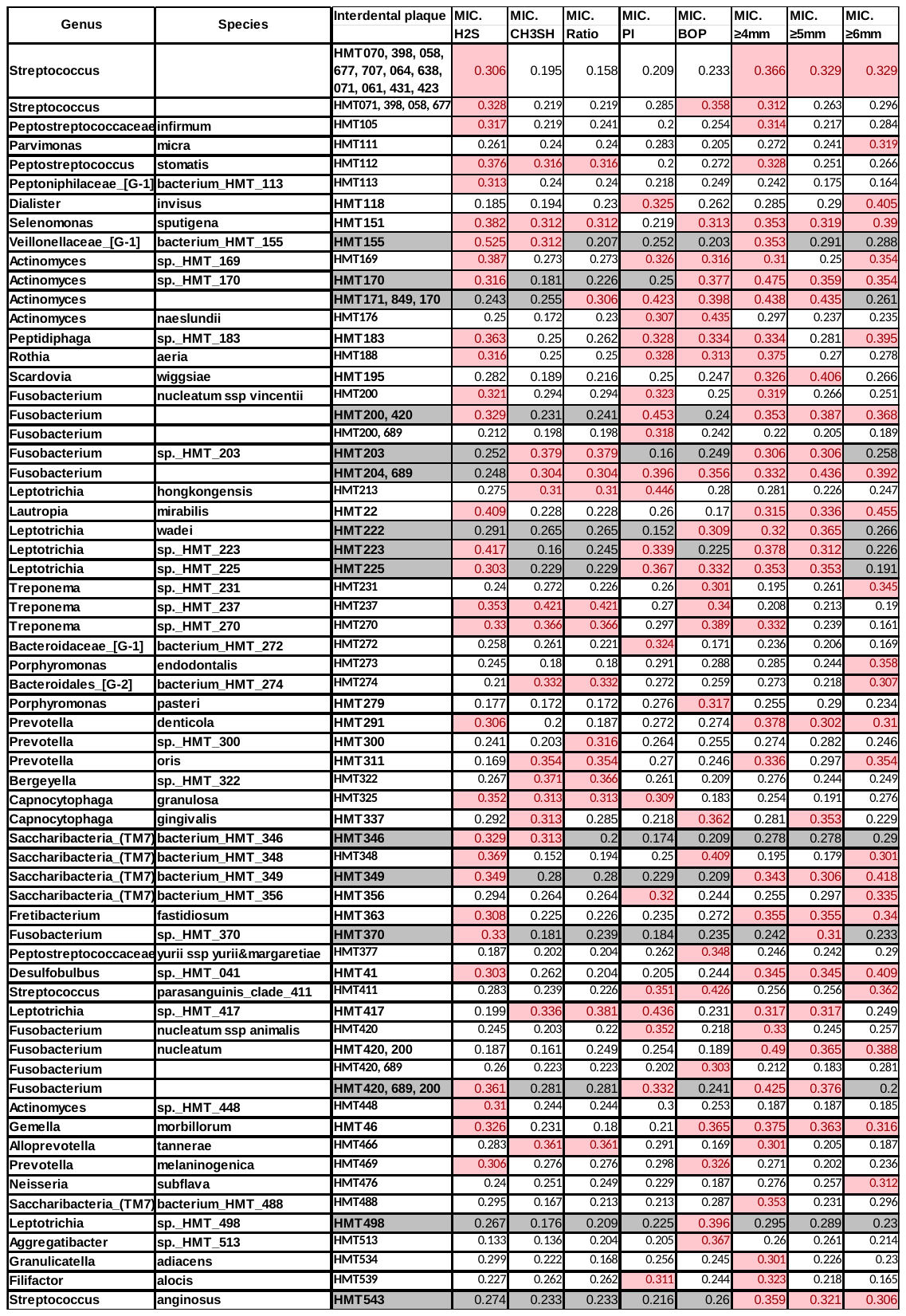

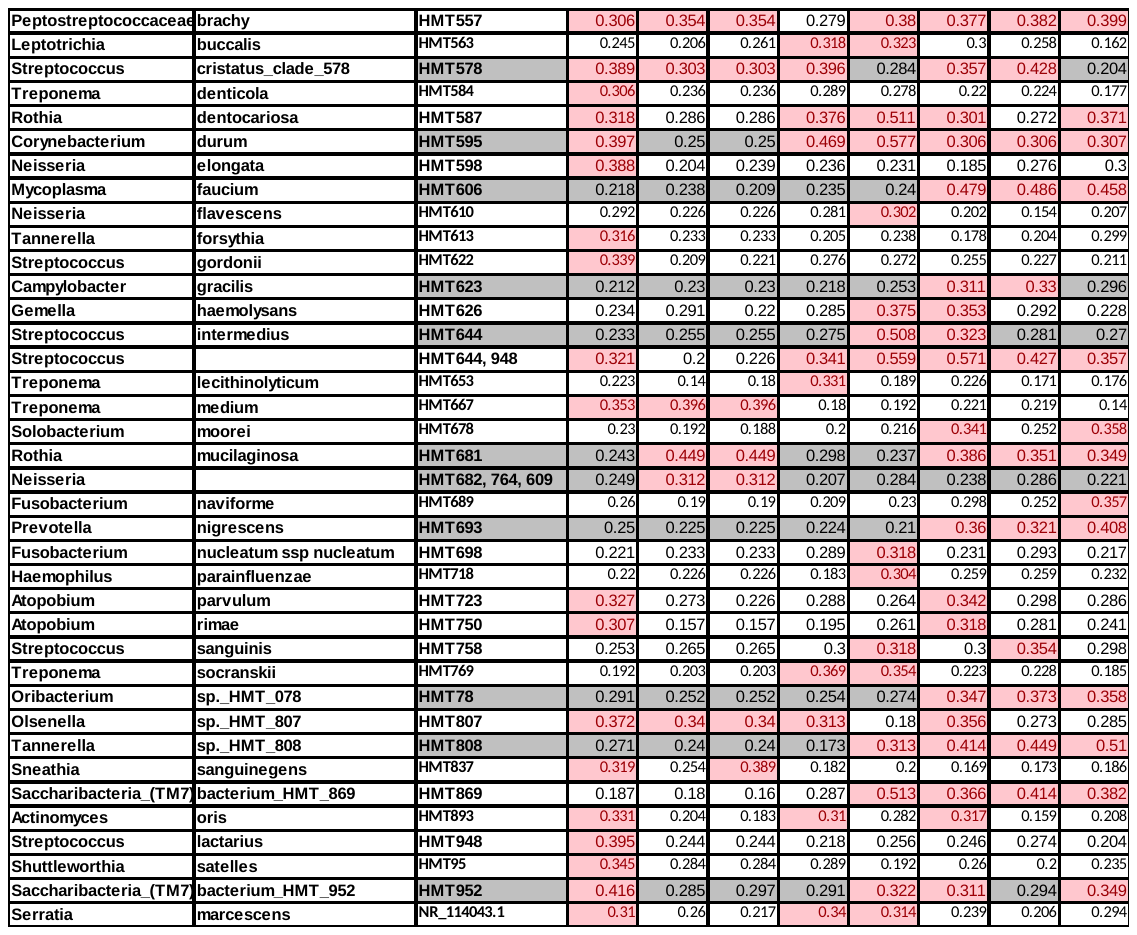

**Supplementary Table S9. Table listing all comparisons for the taxa within the genus *Streptococcus* spp. that showed significant differences as displayed in Figure 4. I=interdental plaque, s=subgingival plaque, t=tongue, h=low plaque health, hp=high plaque health, g=gingivitis, p=periodontitis.**

| Taxa | Tukey's multiple comparisons test | Mean Diff. | 95% CI of diff. | Adjusted P Value |
| --- | --- | --- | --- | --- |
| Streptococcus 073, 066 | ih vs. tg | -0.05863 | -0.09707 to -0.02018 | 0.0033 |
|  | ihp vs. tg | -0.05871 | -0.1032 to -0.01419 | 0.0116 |
|  | sg vs. tg | -0.05808 | -0.1028 to -0.01331 | 0.0128 |
|  | shp vs. tg | -0.05778 | -0.1083 to -0.007287 | 0.0269 |
|  | sp vs. tg | -0.05533 | -0.09215 to -0.01850 | 0.0037 |
| Streptococcus 073, 074, 066 | ig vs. tg | -0.01134 | -0.01745 to -0.005230 | 0.0004 |
|  | ih vs. tg | -0.01137 | -0.01749 to -0.005239 | 0.0008 |
|  | ihp vs. tg | -0.01118 | -0.01749 to -0.004868 | 0.002 |
|  | ip vs. tg | -0.01153 | -0.01756 to -0.005502 | 0.0003 |
|  | sh vs. tg | -0.00991 | -0.01872 to -0.001100 | 0.0333 |
|  | shp vs. tg | -0.01047 | -0.01582 to -0.005115 | 0.0017 |
|  | sp vs. tg | -0.00967 | -0.01769 to -0.001656 | 0.0163 |
| S. cristatus | ig vs. tg | 0.04663 | 0.01314 to 0.08011 | 0.0049 |
|  | ig vs. th | 0.05224 | 0.006614 to 0.09787 | 0.0252 |
|  | ig vs. thp | 0.04577 | 0.006025 to 0.08551 | 0.023 |
|  | ihp vs. tg | 0.07768 | 0.01139 to 0.1440 | 0.0223 |
|  | ihp vs. th | 0.0833 | 0.009976 to 0.1566 | 0.0262 |
|  | ihp vs. thp | 0.07682 | 0.01955 to 0.1341 | 0.0106 |
| S. intermedius/lactarius | ig vs. tg | 0.005712 | 0.0004540 to 0.01097 | 0.0297 |
|  | ihp vs. tg | 0.01154 | 0.0006277 to 0.02246 | 0.0379 |
| S. lactarius | ig vs. thp | 0.01664 | 0.002154 to 0.03113 | 0.0234 |
| S. parasanguinis II | ig vs. tg | -0.1507 | -0.2600 to -0.04132 | 0.0053 |
|  | ig vs. th | -0.2478 | -0.4881 to -0.007633 | 0.0429 |
|  | ih vs. tg | -0.1416 | -0.2720 to -0.01123 | 0.0312 |
|  | ih vs. th | -0.2388 | -0.4553 to -0.02227 | 0.0306 |
|  | ihp vs. tg | -0.1527 | -0.2826 to -0.02282 | 0.0219 |
|  | ip vs. th | -0.2153 | -0.3989 to -0.03178 | 0.0221 |
|  | sg vs. tg | -0.129 | -0.2533 to -0.004694 | 0.0417 |
|  | shp vs. tg | -0.1537 | -0.2785 to -0.02892 | 0.0188 |
| S. parasanguinis II/ HMT 057 | ig vs. tg | -0.0025 | -0.004472 to -0.0005186 | 0.0104 |
|  | ig vs. thp | -0.00331 | -0.005236 to -0.001382 | 0.0018 |
|  | ih vs. tg | -0.00255 | -0.004918 to -0.0001863 | 0.0325 |
|  | ih vs. thp | -0.00337 | -0.005425 to -0.001307 | 0.0025 |
|  | ihp vs. thp | -0.00338 | -0.005829 to -0.0009377 | 0.0089 |
|  | ip vs. tg | -0.0025 | -0.004542 to -0.0004643 | 0.0127 |
|  | ip vs. thp | -0.00332 | -0.005295 to -0.001339 | 0.0021 |
|  | sg vs. thp | -0.00337 | -0.005817 to -0.0009141 | 0.0093 |
|  | shp vs. thp | -0.00338 | -0.006213 to -0.0005400 | 0.0222 |
|  | sp vs. tg | -0.00245 | -0.004732 to -0.0001696 | 0.0332 |
|  | sp vs. thp | -0.00327 | -0.005422 to -0.001108 | 0.0042 |
| S. perioris/lactarius | ig vs. tg | -0.02923 | -0.05038 to -0.008086 | 0.0052 |
|  | ih vs. tg | -0.02977 | -0.04942 to -0.01013 | 0.0035 |
|  | ihp vs. tg | -0.03037 | -0.06032 to -0.0004196 | 0.0467 |
|  | sg vs. tg | -0.0302 | -0.05871 to -0.001700 | 0.0375 |
|  | shp vs. tg | -0.03153 | -0.06301 to -5.445e-005 | 0.0496 |
| S. sanguinis | ig vs. tg | 0.08708 | 0.002924 to 0.1712 | 0.0405 |
|  | ig vs. th | 0.08748 | 0.01693 to 0.1580 | 0.0164 |
|  | ig vs. thp | 0.08231 | 0.01468 to 0.1499 | 0.0167 |
|  | ih vs. tg | 0.06006 | 0.003704 to 0.1164 | 0.0348 |
|  | ih vs. thp | 0.05529 | 0.004301 to 0.1063 | 0.0324 |
| Streptococcus HMT 74 | ig vs. tg | -0.1278 | -0.2363 to -0.01937 | 0.0171 |
|  | ih vs. tg | -0.1308 | -0.2501 to -0.01137 | 0.0297 |
|  | ihp vs. tg | -0.1296 | -0.2447 to -0.01447 | 0.0275 |
|  | ip vs. tg | -0.1301 | -0.2376 to -0.02265 | 0.0141 |
|  | shp vs. tg | -0.1211 | -0.2403 to -0.001974 | 0.0465 |
| Streptococcus 070, 398, 058, 677, 707, 064, 638, 071, 061, 431, 423 | ig vs. tg | 0.317 | 0.08279 to 0.5512 | 0.0061 |
|  | ig vs. th | 0.382 | 0.07581 to 0.6881 | 0.0158 |
|  | ig vs. thp | 0.3648 | 0.02254 to 0.7071 | 0.0356 |
|  | ih vs. tg | 0.3606 | 0.03232 to 0.6889 | 0.0292 |
|  | ih vs. th | 0.4256 | 0.1326 to 0.7186 | 0.0067 |
|  | ih vs. thp | 0.4084 | 0.05246 to 0.7644 | 0.0235 |
|  | ih vs. tp | 0.3925 | 0.06900 to 0.7161 | 0.0185 |
|  | sg vs. th | 0.4636 | 0.03647 to 0.8907 | 0.0332 |
|  | shp vs. tg | 0.343 | 0.003616 to 0.6823 | 0.0477 |
| Streptococcus 066, 721 | ig vs. tg | -0.09983 | -0.1760 to -0.02364 | 0.0079 |
|  | ih vs. tg | -0.09993 | -0.1805 to -0.01941 | 0.0136 |
|  | ihp vs. tg | -0.1001 | -0.1971 to -0.003035 | 0.043 |
|  | ip vs. tg | -0.09165 | -0.1716 to -0.01170 | 0.0207 |
|  | sg vs. tg | -0.09626 | -0.1910 to -0.001503 | 0.0463 |
| Streptococcus 071, 423, 058, 707, 734, 061, 070, 431, 677, 398, 064, 638, 578 | ihp vs. tg | 0.001567 | 0.0002948 to 0.002840 | 0.017 |
|  | ihp vs. th | 0.001551 | 7.171e-005 to 0.003030 | 0.0396 |
|  | ihp vs. thp | 0.001535 | 6.979e-005 to 0.003000 | 0.0397 |
|  | ihp vs. tp | 0.001405 | 0.0001763 to 0.002633 | 0.0253 |

**Supplementary Table S10. Table listing all comparisons for the taxa within the genus *Fusobacterium* spp. that showed significant differences as displayed in Figure 4. I=interdental plaque, s=subgingival plaque, t=tongue, h=low plaque health, hp=high plaque health, g=gingivitis, p=periodontitis.**

| Taxa | Tukey's multiple comparisons test | Mean Diff. | 95% CI of diff. | Adjusted P Value |
| --- | --- | --- | --- | --- |
| Fusobacterium nucleatum subsp. animalis | ig vs. tg | 0.3069 | 0.01217 to 0.6016 | 0.039 |
|  | tg vs. shp | -0.3589 | -0.6960 to -0.02173 | 0.0377 |
| Fusobacterium nucleatum subsp. animalis/vincentii, Fusobacterium naviforme | tg vs. ih | -0.01827 | -0.03274 to -0.003792 | 0.0122 |
|  | ih vs. thp | 0.01847 | 0.0007517 to 0.03618 | 0.0401 |
| Fusobacterium nucleatum subsp. nucleatum | ig vs. th | 0.006456 | 0.0003349 to 0.01258 | 0.0384 |
|  | ig vs. thp | 0.006594 | 0.001048 to 0.01214 | 0.0191 |
| Fusobacterium nucleatum subsp. vincentii | ig vs. tg | 0.2927 | 0.08237 to 0.5030 | 0.0049 |
|  | sg vs. tg | 0.3954 | 0.01386 to 0.7769 | 0.042 |
|  | tg vs. ihp | -0.2888 | -0.5631 to -0.01459 | 0.0387 |
|  | tg vs. ip | -0.2847 | -0.5534 to -0.01600 | 0.0349 |
| Fusobacterium nucleatum subsp. vincentii/nucleatum/animalis, F. naviforme | tg vs. ihp | -0.00271 | -0.005337 to -8.963e-005 | 0.0424 |
| Fusobacterium nucleatum subsp. vincentii/animalis | ig vs. tg | 0.04342 | 0.01401 to 0.07283 | 0.0031 |
|  | ig vs. th | 0.04288 | 0.004987 to 0.08077 | 0.0268 |
| Fusobacterium periodonticum | ig vs. tg | -0.8506 | -1.039 to -0.6617 | <0.0001 |
|  | ig vs. th | -0.7672 | -1.254 to -0.2805 | 0.0041 |
|  | ig vs. thp | -0.8656 | -1.103 to -0.6281 | <0.0001 |
|  | ig vs. tp | -0.6891 | -1.161 to -0.2172 | 0.0065 |
|  | sg vs. tg | -0.827 | -1.039 to -0.6151 | <0.0001 |
|  | sg vs. th | -0.7436 | -1.253 to -0.2343 | 0.0065 |
|  | sg vs. thp | -0.842 | -1.103 to -0.5811 | <0.0001 |
|  | sg vs. tp | -0.6655 | -1.146 to -0.1848 | 0.0088 |
|  | tg vs. ih | 0.8574 | 0.7362 to 0.9787 | <0.0001 |
|  | tg vs. sh | 0.841 | 0.5644 to 1.118 | 0.0007 |
|  | tg vs. ihp | 0.8571 | 0.7143 to 0.9998 | <0.0001 |
|  | tg vs. shp | 0.8401 | 0.6637 to 1.017 | <0.0001 |
|  | tg vs. ip | 0.8498 | 0.6492 to 1.050 | <0.0001 |
|  | tg vs. sp | 0.8552 | 0.7322 to 0.9782 | <0.0001 |
|  | ih vs. th | -0.7741 | -1.260 to -0.2879 | 0.0039 |
|  | ih vs. thp | -0.8725 | -1.105 to -0.6396 | <0.0001 |
|  | ih vs. tp | -0.696 | -1.186 to -0.2060 | 0.0076 |
|  | sh vs. thp | -0.8561 | -1.303 to -0.4091 | 0.0046 |
|  | sh vs. tp | -0.6796 | -1.324 to -0.03482 | 0.0417 |
|  | th vs. ihp | 0.7737 | 0.2619 to 1.286 | 0.0053 |
|  | th vs. shp | 0.7568 | 0.1723 to 1.341 | 0.0147 |
|  | th vs. ip | 0.7665 | 0.3015 to 1.231 | 0.0031 |
|  | th vs. sp | 0.7719 | 0.2862 to 1.258 | 0.0039 |
|  | ihp vs. thp | -0.8721 | -1.143 to -0.6013 | <0.0001 |
|  | ihp vs. tp | -0.6956 | -1.211 to -0.1800 | 0.0102 |
|  | shp vs. thp | -0.8552 | -1.152 to -0.5589 | 0.0002 |
|  | shp vs. tp | -0.6787 | -1.266 to -0.09127 | 0.0257 |
|  | thp vs. ip | 0.8649 | 0.6456 to 1.084 | <0.0001 |
|  | thp vs. sp | 0.8703 | 0.6360 to 1.105 | <0.0001 |
|  | ip vs. tp | -0.6884 | -1.158 to -0.2185 | 0.0064 |
|  | sp vs. tp | -0.6938 | -1.184 to -0.2031 | 0.0078 |

**Supplementary Table S11. Table listing all comparisons for the taxa within the genus *Actinomyces* spp. that showed significant differences as displayed in Figure 4. I=interdental plaque, s=subgingival plaque, t=tongue, h=low plaque health, hp=high plaque health, g=gingivitis, p=periodontitis.**

| Taxa | Tukey's multiple comparisons test | Mean Diff. | 95% CI of diff. | Adjusted P Value |
| --- | --- | --- | --- | --- |
| Actinomyces HMT169 | sg vs. tg | 0.03664 | 0.01487 to 0.05840 | 0.0028 |
|  | sg vs. th | 0.03743 | 0.01300 to 0.06185 | 0.0049 |
|  | sg vs. thp | 0.0368 | 0.01531 to 0.05830 | 0.0025 |
|  | sg vs. tp | 0.03473 | 0.01096 to 0.05849 | 0.0064 |
|  | tg vs. ihp | -0.06017 | -0.1114 to -0.008937 | 0.022 |
|  | tg vs. ip | -0.2502 | -0.4802 to -0.02008 | 0.0295 |
|  | th vs. ihp | -0.06097 | -0.1173 to -0.004673 | 0.0336 |
|  | ihp vs. thp | 0.06034 | 0.003251 to 0.1174 | 0.038 |
|  | ihp vs. tp | 0.05827 | 0.002507 to 0.1140 | 0.0403 |
|  | thp vs. ip | -0.2503 | -0.4878 to -0.01282 | 0.0378 |
| Actinomyces HMT172 | ig vs. tg | -0.2235 | -0.4365 to -0.01054 | 0.0371 |
|  | tg vs. shp | 0.2219 | 0.03960 to 0.4042 | 0.0199 |
|  | tg vs. ip | 0.224 | 0.01301 to 0.4349 | 0.0344 |
|  | ihp vs. thp | -0.194 | -0.3677 to -0.02025 | 0.0287 |
|  | thp vs. ip | 0.1923 | 0.008093 to 0.3765 | 0.0398 |
| Actinomyces naeslundii | tg vs. ihp | -0.2346 | -0.4339 to -0.03526 | 0.0218 |
|  | tg vs. shp | -0.1826 | -0.2899 to -0.07526 | 0.0035 |
|  | th vs. ihp | -0.2352 | -0.4546 to -0.01577 | 0.0354 |
|  | th vs. shp | -0.1832 | -0.3004 to -0.06594 | 0.0056 |
|  | ihp vs. thp | 0.2351 | 0.02022 to 0.4500 | 0.0319 |
|  | ihp vs. tp | 0.233 | 0.01163 to 0.4545 | 0.0388 |
|  | shp vs. thp | 0.1831 | 0.07057 to 0.2956 | 0.0045 |
|  | shp vs. tp | 0.181 | 0.06193 to 0.3001 | 0.0065 |
| Actinomyces HMT180 | ig vs. tg | -0.361 | -0.5841 to -0.1380 | 0.0014 |
|  | ig vs. th | -0.3175 | -0.6271 to -0.007819 | 0.0443 |
|  | ig vs. tp | -0.2946 | -0.5889 to -0.0002227 | 0.0498 |
|  | tg vs. ih | 0.3811 | 0.1927 to 0.5695 | 0.0004 |
|  | tg vs. ihp | 0.4137 | 0.1272 to 0.7003 | 0.0069 |
|  | tg vs. ip | 0.3686 | 0.1470 to 0.5903 | 0.0011 |
|  | th vs. ihp | 0.3702 | 0.09682 to 0.6435 | 0.01 |
|  | th vs. ip | 0.3251 | 0.03475 to 0.6154 | 0.0283 |
|  | ihp vs. tp | -0.3473 | -0.6426 to -0.05198 | 0.0218 |
| Actinomyces lingnae [NVP] | ig vs. tg | -0.1383 | -0.2639 to -0.01272 | 0.0271 |
|  | sg vs. tg | -0.1137 | -0.2206 to -0.006924 | 0.0366 |
|  | tg vs. ihp | 0.1403 | 0.004585 to 0.2761 | 0.0425 |
|  | tg vs. ip | 0.1393 | 0.01560 to 0.2630 | 0.0234 |
| Actinomyces HMT448 | ig vs. tg | 0.4166 | 0.01608 to 0.8171 | 0.0392 |
| Actinomyces meyeri | tg vs. ih | -0.0165 | -0.03224 to -0.0007677 | 0.0382 |
| Actinomyces massiliensis | tg vs. ihp | -0.05226 | -0.09211 to -0.01240 | 0.012 |
|  | sh vs. ihp | -0.03745 | -0.07403 to -0.0008613 | 0.0462 |
|  | th vs. ihp | -0.05229 | -0.09664 to -0.007950 | 0.0215 |
|  | ihp vs. thp | 0.05229 | 0.009199 to 0.09539 | 0.0184 |
|  | ihp vs. tp | 0.05229 | 0.007950 to 0.09664 | 0.0215 |
| Actinomyces graevenitzii | ig vs. tg | -0.1005 | -0.1955 to -0.005444 | 0.0354 |
|  | tg vs. ih | 0.09723 | 0.001042 to 0.1934 | 0.0471 |
|  | tg vs. ip | 0.1005 | 0.005413 to 0.1956 | 0.0354 |

**Supplementary Table S12. Table listing all comparisons for the taxa within the genus *Prevotella* spp. that showed significant differences as displayed in Figure 4. I=interdental plaque, s=subgingival plaque, t=tongue, h=low plaque health, hp=high plaque health, g=gingivitis, p=periodontitis.**

| Taxa | Tukey's multiple comparisons test | Mean Diff. | 95% CI of diff. | Adjusted P Value |
| --- | --- | --- | --- | --- |
| P. oris | tg vs. ih | -0.4805 | -0.7063 to -0.2547 | 0.0003 |
|  | tg vs. shp | -0.4012 | -0.7197 to -0.08280 | 0.0168 |
|  | ih vs. th | 0.4775 | 0.2127 to 0.7423 | 0.0018 |
|  | ih vs. thp | 0.4804 | 0.2235 to 0.7372 | 0.001 |
|  | ih vs. tp | 0.4777 | 0.2218 to 0.7336 | 0.0015 |
|  | th vs. shp | -0.3983 | -0.7502 to -0.04637 | 0.0283 |
|  | shp vs. thp | 0.4011 | 0.05881 to 0.7434 | 0.024 |
|  | shp vs. tp | 0.3985 | 0.04805 to 0.7489 | 0.0277 |
| P. melaninogenica | ig vs. tg | -0.4746 | -0.8195 to -0.1297 | 0.0054 |
|  | ig vs. tp | -0.5926 | -0.9866 to -0.1987 | 0.0054 |
|  | sg vs. thp | -0.3453 | -0.6132 to -0.07745 | 0.0132 |
|  | sg vs. tp | -0.5874 | -1.096 to -0.07912 | 0.024 |
|  | tg vs. ih | 0.5657 | 0.2784 to 0.8530 | 0.0005 |
|  | tg vs. shp | 0.5781 | 0.2158 to 0.9405 | 0.005 |
|  | tg vs. ip | 0.4778 | 0.1527 to 0.8030 | 0.0032 |
|  | ih vs. thp | -0.4417 | -0.6557 to -0.2277 | 0.0005 |
|  | ih vs. tp | -0.6837 | -0.8831 to -0.4843 | <0.0001 |
|  | th vs. ip | 0.3362 | 0.07710 to 0.5953 | 0.0127 |
|  | ihp vs. tp | -0.6057 | -1.011 to -0.2007 | 0.0056 |
|  | shp vs. thp | -0.4542 | -0.7354 to -0.1729 | 0.0047 |
|  | shp vs. tp | -0.6962 | -0.9024 to -0.4899 | <0.0001 |
|  | thp vs. ip | 0.3538 | 0.09381 to 0.6139 | 0.0084 |
|  | ip vs. tp | -0.5959 | -0.9284 to -0.2633 | 0.0019 |
|  | sp vs. tp | -0.5415 | -0.8498 to -0.2332 | 0.0021 |
| P. nigrescens | tg vs. ih | -0.07962 | -0.1483 to -0.01093 | 0.021 |
|  | tg vs. shp | -0.2561 | -0.4848 to -0.02748 | 0.0297 |
|  | th vs. shp | -0.2588 | -0.5095 to -0.008108 | 0.0433 |
|  | shp vs. thp | 0.2581 | 0.01190 to 0.5043 | 0.0404 |
| P. pallens | ig vs. tg | -0.01299 | -0.02542 to -0.0005601 | 0.0381 |
|  | tg vs. sp | 0.01376 | 0.002830 to 0.02469 | 0.0124 |

**Supplementary Table S13. Table listing all comparisons for the taxa within the family Porphyromonadaceae that showed significant differences as displayed in Figure 4. I=interdental plaque, s=subgingival plaque, t=tongue, h=low plaque health, hp=high plaque health, g=gingivitis, p=periodontitis.**

| Taxa | Tukey's multiple comparisons test | Mean Diff. | 95% CI of diff. | Adjusted P Value |
| --- | --- | --- | --- | --- |
| Porphyromonas pasteri | ig vs. tg | -0.558 | -1.031 to -0.08450 | 0.0171 |
|  | tg vs. sh | 0.6724 | 0.1243 to 1.221 | 0.0244 |
|  | tg vs. sp | 0.6411 | 0.1123 to 1.170 | 0.0158 |
|  | ih vs. thp | -0.5386 | -1.034 to -0.04311 | 0.0319 |
|  | thp vs. sp | 0.6321 | 0.2069 to 1.057 | 0.0048 |
|  | sp vs. tp | -0.6196 | -1.209 to -0.03050 | 0.039 |
| Tannerella forsythia | tg vs. sp | -0.4736 | -0.9191 to -0.02814 | 0.0353 |

**Supplementary Table S14. Table listing all comparisons for the taxa within the genus *Saccharibacteria* (TM7) spp. that showed significant differences as displayed in Figure 4. I=interdental plaque, s=subgingival plaque, t=tongue, h=low plaque health, hp=high plaque health, g=gingivitis, p=periodontitis.**

| Taxa | Tukey's multiple comparisons test | Mean Diff. | 95% CI of diff. | Adjusted P Value |
| --- | --- | --- | --- | --- |
| **Saccharibacteria HMT346** | **ig vs. tg** | **0.2209** | **0.01345 to 0.4284** | **0.0338** |
| **Saccharibacteria HMT352** | **ig vs. tg** | **-0.9199** | **-1.135 to -0.7053** | **<0.0001** |
|  | **ig vs. th** | **-0.8015** | **-1.304 to -0.2988** | **0.0038** |
|  | **ig vs. thp** | **-0.9645** | **-1.097 to -0.8324** | **<0.0001** |
|  | **ig vs. tp** | **-0.7305** | **-1.314 to -0.1466** | **0.0156** |
|  | **sg vs. tg** | **-0.9407** | **-1.015 to -0.8668** | **<0.0001** |
|  | **sg vs. th** | **-0.8222** | **-1.366 to -0.2786** | **0.0053** |
|  | **sg vs. thp** | **-0.9853** | **-1.014 to -0.9563** | **<0.0001** |
|  | **sg vs. tp** | **-0.7513** | **-1.400 to -0.1021** | **0.0238** |
|  | **tg vs. ih** | **0.9196** | **0.6102 to 1.229** | **<0.0001** |
|  | **tg vs. sh** | **0.9112** | **0.7276 to 1.095** | **0.0002** |
|  | **tg vs. ihp** | **0.9425** | **0.8698 to 1.015** | **<0.0001** |
|  | **tg vs. shp** | **0.9454** | **0.8653 to 1.025** | **<0.0001** |
|  | **tg vs. ip** | **0.9382** | **0.7398 to 1.137** | **<0.0001** |
|  | **tg vs. sp** | **0.9451** | **0.7052 to 1.185** | **<0.0001** |
|  | **ih vs. th** | **-0.8012** | **-1.313 to -0.2893** | **0.0043** |
|  | **ih vs. thp** | **-0.9643** | **-1.054 to -0.8745** | **<0.0001** |
|  | **ih vs. tp** | **-0.7302** | **-1.342 to -0.1183** | **0.0202** |
|  | **sh vs. thp** | **-0.9559** | **-1.109 to -0.8026** | **<0.0001** |
|  | **th vs. ihp** | **0.8241** | **0.2802 to 1.368** | **0.0052** |
|  | **th vs. shp** | **0.8269** | **0.1931 to 1.461** | **0.0141** |
|  | **th vs. ip** | **0.8197** | **0.3271 to 1.312** | **0.003** |
|  | **th vs. sp** | **0.8266** | **0.3086 to 1.345** | **0.0038** |
|  | **ihp vs. thp** | **-0.9871** | **-1.026 to -0.9485** | **<0.0001** |
|  | **ihp vs. tp** | **-0.7531** | **-1.404 to -0.1023** | **0.0238** |
|  | **shp vs. thp** | **-0.99** | **-1.005 to -0.9754** | **<0.0001** |
|  | **thp vs. ip** | **0.9828** | **0.9364 to 1.029** | **<0.0001** |
|  | **thp vs. sp** | **0.9897** | **0.9648 to 1.015** | **<0.0001** |
|  | **ip vs. tp** | **-0.7487** | **-1.336 to -0.1613** | **0.0141** |
|  | **sp vs. tp** | **-0.7556** | **-1.375 to -0.1360** | **0.018** |

**Supplementary Table S15. Table listing all comparisons for the taxa within the phyla Spirochaetes and Synergistetes that showed significant differences as displayed in Figure 4. I=interdental plaque, s=subgingival plaque, t=tongue, h=low plaque health, hp=high plaque health, g=gingivitis, p=periodontitis.**

| Taxa | Tukey's multiple comparisons test | Mean Diff. | 95.00% CI of diff. | Adjusted P Value |
| --- | --- | --- | --- | --- |
| Treponema HMT231 | ig vs. th | 0.1469 | 0.01299 to 0.2807 | 0.0315 |
|  | ig vs. thp | 0.1469 | 0.01221 to 0.2815 | 0.0313 |
| Fretibacterium HMT359 | ig vs. th | 0.01419 | 0.002477 to 0.02590 | 0.0186 |
|  | ig vs. tp | 0.01277 | 0.0006607 to 0.02488 | 0.0385 |
| Fretibacterium HMT360 | tg vs. shp | -0.03232 | -0.06405 to -0.0005964 | 0.046 |
|  | tg vs. ip | -0.00822 | -0.01510 to -0.001343 | 0.0154 |
|  | th vs. ip | -0.00822 | -0.01517 to -0.001271 | 0.0212 |
|  | ip vs. tp | 0.008219 | 0.001271 to 0.01517 | 0.0212 |
| Fretibacterium fastidiosum | sg vs. thp | 0.03894 | 0.008700 to 0.06918 | 0.0133 |
| Treponema socranskii | sg vs. thp | 0.4148 | 0.009517 to 0.8201 | 0.0446 |
|  | tg vs. shp | -0.4154 | -0.7226 to -0.1082 | 0.0118 |
|  | tg vs. sp | -0.2363 | -0.4093 to -0.06327 | 0.0072 |
|  | shp vs. thp | 0.5077 | 0.1730 to 0.8425 | 0.0065 |
|  | shp vs. tp | 0.4201 | 0.05700 to 0.7832 | 0.0255 |

**Supplementary Table S16. Table listing all comparisons for the taxa within the genus *Veillonella* spp. that showed significant differences as displayed in Figure 4. I=interdental plaque, s=subgingival plaque, t=tongue, h=low plaque health, hp=high plaque health, g=gingivitis, p=periodontitis.**

| Taxa | Tukey's multiple comparisons test | Mean Diff. | 95% CI of diff. | Adjusted P Value |
| --- | --- | --- | --- | --- |
| V. parvula | sh vs. ihp | 0.177 | 0.03593 to 0.3181 | 0.0225 |
| V. atypica | ig vs. tg | -0.1923 | -0.3522 to -0.03239 | 0.0148 |
|  | tg vs. ih | 0.192 | 0.03265 to 0.3515 | 0.0165 |
|  | tg vs. ip | 0.1857 | 0.03395 to 0.3375 | 0.0131 |
|  | tg vs. sp | 0.184 | 0.01547 to 0.3525 | 0.0302 |
